# Autism Sensory Profiles Predict Stimulus-Evoked Insula Connectivity

**DOI:** 10.64898/2026.04.29.26352062

**Authors:** Zachary Jacokes, Stefen Beeler-Duden, Sophie Lawson, Jeff Eilbott, Mirella Dapretto, Natalia Kleinhans, James McPartland, Allison Jack, Gregory Wallace, Denis Sukhodolsky, Sarah Jane Webb, John Darrell Van Horn, Kevin Pelphrey, the GENDAAR Research Consortium

## Abstract

Sensory processing is a common target in autism spectrum disorder (ASD) research, yet the latent structure of sensory experience is disputed. Researchers frequently explore the presence of “subtypes” to categorize sensory heterogeneity, but such discrete models can fail to capture the intrinsic geometry of phenotypic data. In this study, we aim to characterize heterogeneous sensory profiles in ASD and explore if the same characterization can describe neurobiological function. First, we apply unsupervised spectral manifold dimensionality reduction to item-level Sensory Profile data from a large cohort of autistic participants (*n*=223) to compare categorical subtyping against continuous models. The behavioral results reveal unstable and irreproducible subtyping solutions; instead, sensory processing differences are best characterized as a continuous, non-linear manifold of sensory severity.

To determine the neurobiological relevance of this sensory gradient, we employed voxel-wise linear mixed-effects modeling of insula-seeded functional connectivity (*n*=63). We demonstrate that sensory severity predicts a significant decoupling between the insula and sensorimotor cortices during externally driven stimulation involving motion stimuli, but not during resting state. This finding supports the interpretation that sensory-related neural hypoconnectivity is context-dependent and not reflective of intrinsic traits. Further, we identify a significant sex-by-sensory gradient interaction, indicating heightened sensitivity of connectivity patterns to sensory severity in autistic males. These findings indicate that sensory atypicality in ASD points toward a continuous regulatory manifold linked to disrupted social-sensory integration.

## 1. Introduction

Autism spectrum disorder (ASD) is phenotypically and neurobiologically heterogeneous, involving differences in social behavior, language, and sensory processing^1–3^. Yet much of the ASD research has been focused on how autistic individuals differ from their non-autistic counterparts^4,5^. While comparisons between ASD and non-ASD individuals may improve detection and diagnosis, there is growing interest in within-ASD heterogeneity and data-driven subtyping^6,7^. Elucidating the heterogeneity of ASD could indeed address some of the limitations of traditional case-control studies^5^.

Recent advances in machine learning have enabled categorization of highly dimensional behavioral, cognitive, and neuroimaging data. Importantly, these methods move beyond discrete classification systems and allow for richly detailed within-group examinations using continuous data commonly found in neuropsychological assessments. These categorizations may move ASD interventions towards a personalized medicine approach, with clinical decisions tailored to individual needs. Further, ASD diagnostic analyses have identified a stark four-to-one sex-wise diagnostic disparity, where boys are far more likely to be diagnosed with ASD than girls^8^. This disparity implies the presence of genetic, biological or environmental factors that may affect the development of ASD in boys and girls differently. Some hypotheses proposed to explain this disparity center on hormonal differences, genetic and epigenetic factors, and neurobiological underpinnings^9–12^. Sex differences may reflect different mappings between behavioral phenotypes and neural systems, and any model of heterogeneity that ignores sex may conflate distinct regulatory trajectories. Gaining a more holistic understanding of sex-wise differences in ASD is vitally important for personalized treatment plans for affected individuals.

This work aims to explore heterogeneity within ASD specifically through the lens of sensory response. Both phenotypic experience and functional neurobiology are examined to establish a connection between behavior and neurobiology in ASD. Importantly, this analysis attempts to assess whether ASD-specific sensory experience manifests as static traits, or if it is state-dependent. Crucially, we did not model task-evoked activation contrasts; rather, we examined how sensory severity predicts large-scale patterns of functional connectivity across contexts. We assess this using self-reported Sensory Profile data and task-based fMRI, particularly focusing on the insula, a region heavily involved in salience and sensory integration^13^.

### 1.1. Sensory Processing in ASD

Sensory differences are a core feature of ASD and warrant deep exploration due to the early developmental onset and relevance across domains, including motor and emotion processing^14,15^. Prior literature is divided on whether these differences reflect distinct subtypes or a more subtle gradient of atypical sensory processing^16^. Thye and colleagues explore the domains in which autistic individuals express sensory differences (auditory and tactile processing, olfaction and gustation, and integration therein), highlighting the heterogeneity of sensory experiences in ASD^17^. Marco and colleagues report on the neurophysiology of sensory processing in ASD, suggesting that multisensory integration in brain regions such as the prefrontal cortex and the temporal lobe may be impaired in autistic individuals^18^. Sensory processing differences persist throughout adolescence in autistic individuals, though the degree of these differences appears to decrease over time^19^.

Beyond the established heterogeneity, biological sex remains a primary factor in ASD sensory neurobiology. Recent studies suggest that autistic females may exhibit different sensory phenotypes than their male counterparts, characterized by higher levels of sensory sensitivity but better compensatory behavioral strategies^20–22^. Prior work has identified neurobiological sex differences in the structural and functional connectivity of the salience network, particularly the insula, suggesting that the neural correlates of sensory atypicality may not be uniform across sexes^23^. Understanding these divergent paths is critical to ensure that sensory-based neurological profiles are not inadvertently biased toward male-typical presentations.

### 1.2. Role of the Insula in Sensory Processing

The insula can be subdivided into three functional subregions: the dorsal anterior-to-middle insula (dAI), the ventral anterior insula (vAI), and the posterior insula (PI)^24,25^. The insula has been identified as a salience-integration-action hub, implicated in network switching between the default mode and salience networks, interoceptive processes, and motor system coupling^16,26^.

The dAI has been implicated in cognitive control and salience detection. It is functionally connected to the dorsal anterior cingulate cortex and the prefrontal cortex via the salience and frontoparietal control networks. It is responsible for detecting salient events, coordinating network switching, and supporting decision-making^24,26^. The vAI subregion is responsible for affective and social-emotional processing. It is functionally connected to the amygdala, ventral striatum, orbitofrontal cortex, and limbic regions. It is implicated in emotion processing, importantly including empathy and social emotions. It also links interoceptive signals with affective experience^24,26^. The PI subregion is primarily responsible for sensorimotor and interoceptive processing. It has functional associations with sensorimotor, somatosensory, auditory cortex, and posterior temporal regions. It helps to process pain, temperature, touch, and other visceral sensations; it essentially represents the physiological condition of the body^24,26^.

Prior research into the insula has revealed linkages to ASD. Both hypo- and hyper-connectivity to and from the insula subregions have been shown to be associated with ASD severity^27^. The insula serves as a bridge between perception and action, a framework often described as the “action-perception loop”^28,29^. In ASD, this coupling appears disrupted. Rather than viewing sensory symptoms as isolated perceptual deficits, emerging models suggest they may reflect an inability to integrate sensory input with motor readiness^30,31^. If the PI cannot efficiently relay interoceptive or somatosensory signals to the primary motor cortex, the individual may struggle to calibrate their behavioral responses to environmental stimuli. The present analysis investigates whether this insula-sensorimotor coupling is modulated by sensory severity, particularly when the sensory input requires social interpretation.

### 1.3. Social-Sensory Integration and Biological Motion

Resting state fMRI provides a window into intrinsic brain functioning, but it may fail to capture the dynamic sensory challenges faced by autistic individuals in daily life. In ASD, sensory noise is frequently most overwhelming when embedded within socially meaningful contexts, which require the simultaneous integration of multi-modal sensory input and social cognition. To probe this, we investigated the “Biopoint” task, a paradigm originally developed by Klin and colleagues (and refined by Kaiser and colleagues) that uses point-light displays to depict biological motion^32,33^. Biological motion is a foundational social-sensory stimulus that requires the brain to interpret human intent from basic visual kinetics. By comparing connectivity during this task to resting state, we can determine if ASD-specific sensory-related neurobiology is defined by static traits or social stimulus-dependent sensory integration.

### 1.4. Sensory Profile

A substantial portion of sensory-processing research in ASD relies on caregiver-report questionnaires, which include the Sensory Profile and Sensory Profile–2 (SP), and the Short Sensory Profile (short-SP). These were developed within the occupational therapy domain^34,35^. These instruments were explicitly designed to support clinical reasoning in occupational therapeutic practice by characterizing patterns of sensory-related behaviors in everyday contexts rather than isolating specific sensory or perceptual mechanisms. Accordingly, these sensory measures are organized around sensory “sections” (e.g., auditory, tactile, movement) and behavioral “quadrants” reflecting Dunn’s sensory processing framework, which integrates low registration, sensation seeking, sensory sensitivity, and sensation avoiding with regulatory and contextual demands.

Early comparative work using the SP questionnaires demonstrates that most young children with ASD exhibit clinically significant sensory differences relative to non-autistic peers, with particularly large group differences in under-responsivity, auditory filtering, and tactile sensitivity^36^. These findings help to establish sensory features as both prevalent and quantifiable at the behavioral level in ASD.

Critically, many of the items in the SP questionnaires intentionally conflate sensory sensitivity and behavioral regulation, which is consistent with their clinical goal of capturing sensory responses in daily life. For example, items often index emotional or behavioral responses to sensory stimulation (such as distress, withdrawal, distractibility, or seeking behaviors) in addition to their perceptual thresholds. As a result, SP-derived scores reflect functional sensory–regulatory patterns rather than separable dimensions of sensory reactivity, modulation, or affective regulation. This design choice is aligned with occupational therapy practice, where sensory processing is conceptualized as inherently embedded within self-regulation, participation, and environmental demands, but it poses challenges for research aiming to isolate sensory encoding from downstream emotional or regulatory processes.

At the same time, the interpretability of SP-derived subscales and total scores depends on whether the instrument measures stable and invariant latent constructs in autistic and non-autistic populations. Williams and colleagues conducted a comprehensive psychometric evaluation of the short-SP in a large sample of autistic individuals and found poor support for the proposed factor structure^37^. Their analyses indicated that short-SP subscales do not consistently map onto stable latent constructs in ASD and that the short-SP total score, although reliable, may reflect substantial multidimensionality rather than a single general sensory factor. Consequently, they cautioned against interpreting short-SP subscale scores and, to a lesser extent, total scores as straightforward indices of distinct sensory processes in autistic participants. Previous neuroimaging studies have found that total scores and summary results from tests such as the Autism Diagnostic Observation Schedule (ADOS) are not as closely associated with structural brain metrics compared to domain and task specific sub-scores^38–40^.

These measurement concerns are particularly relevant for studies that use SP as inputs for person-centered modeling approaches, such as subtyping or latent profile analysis. If the underlying structure of the measure is unstable or non-invariant, identified “subtypes” may partially reflect instrument-specific variance or analytic decisions rather than robust latent sensory phenotypes. This motivates complementary approaches that emphasize item-level structure, dimensional modeling, and validation against neurobiological measures.

### 1.5. Prior Sensory-based Subtyping Analyses

An unresolved question in the sensory autism literature is whether individual differences in sensory processing reflect discrete phenotypes or continuous variation along one or more latent dimensions. In response, a substantial body of work has applied data-centered statistical approaches to sensory questionnaire data in ASD, including latent profile analysis, mixture models, and clustering algorithms.

A widely cited example is the work of Ausderau and colleagues, who applied latent profile transition analysis to a large national caregiver-report dataset spanning early childhood to preadolescence and identified four sensory “subtypes” that appeared relatively stable over a one-year interval^41^. These profiles included patterns interpreted as qualitatively distinct alongside profiles that differed primarily in overall severity. This framework has been influential in motivating sensory phenotyping as a means of parsing heterogeneity in ASD and linking sensory patterns to functional outcomes.

Subsequent studies have extended sensory subtyping to longitudinal and transdiagnostic contexts while highlighting the sensitivity of subtype solutions to modeling choices and measurement characteristics. Dwyer and colleagues used growth mixture modeling to identify longitudinal classes of sensory responsivity that were largely distinguished by overall intensity of sensory difficulties, with one smaller class characterized by increasing difficulties over time. In related work, sensory subtyping approaches have identified subgroups in which autistic and non-autistic individuals group together, suggesting that a substantial proportion of variance in sensory responsivity may be organized along a continuum that spans diagnostic categories rather than forming autism-specific categorical groups^42^.

More recent work in mixed neurodevelopmental samples has reported sensory profiles that cross diagnostic boundaries and align with broader dimensions of self-regulation, emotion dysregulation, anxiety, and attention-deficit/hyperactivity symptoms^43^. These findings raise the possibility that sensory subtypes may reflect shared transdiagnostic mechanisms rather than distinct autism-specific sensory phenotypes. Concurrently, application-oriented studies have demonstrated that established sensory subtype frameworks can show differential associations with motor skills, adaptive behavior, and core autism features, underscoring their clinical appeal while also revealing practical constraints related to subgroup size and stability^44^. In previous work by our group, we applied an experimental approach using a hierarchical random forest clustering method^45^ to group ASD and non-ASD individuals using a mixture of social, behavioral, language, sensory, and structural neuroimaging measures^39^. This approach uncovered two distinct ASD subtypes, mediated by age, and separated by distinct behavioral profiles, with one group displaying high sensory sensitivity values and language performance while the other was characterized by high restrictive and repetitive behaviors.

Across the literature, several methodological themes recur. First, many subtyping solutions appear to be driven largely by overall severity or global atypicality rather than sharply separable qualitative categories. Second, subtype membership and number vary depending on whether analyses use SP totals, subscales, or item-level data, as well as on model selection criteria. Third, psychometric concerns about sensory questionnaires in ASD complicate interpretation of identified profiles as latent sensory phenotypes. Taken together, this body of work motivates approaches that explicitly allow for continuous latent structure and that test whether categorical grouping provides explanatory value beyond a low-dimensional gradient of sensory responsivity, particularly when evaluated against neural systems involved in integrating sensory input with action in socially meaningful contexts.

In the present study, functional magnetic resonance imaging (fMRI) and resulting functional connectivity (FC) are leveraged to explore the neurobiologically-grounded underpinnings of ASD. The gaps in the literature are addressed by moving beyond categorical subtyping to explore sensory processing as a continuous latent dimension. Using unsupervised spectral embedding on item-level Sensory Profile data, this analysis identifies a robust sensory gradient within a large, sex-balanced autistic cohort. This sensory gradient is then validated against neurobiological data using voxel-wise linear mixed-effects models to examine insula-seeded functional connectivity, specifically testing whether individual differences along this sensory gradient modulate the brain’s functional architecture in a context-dependent manner.

## 2. Methods

Data from Wave 1 of the original Autism Centers of Excellence – Gender Exploration of Neurogenetics and Development to Advanced Autism Research (ACE GENDAAR) project were collected across four sites: Yale University, Harvard University/Boston Children’s Hospital, the University of California, Los Angeles, and the University of Washington/Seattle Children’s Research Institute. All procedures were approved by the institutional review boards of participating sites and conducted in accordance with the Declaration of Helsinki. The study received institutional review board (IRB) approvals as follows: Yale as lead institution (IRB: 1206010363; Pelphrey), UCLA (IRB# 10-000387-CR-00008; Bookheimer), Boston Children’s Hospital (IRB-P00004852; Nelson), and with Seattle Children’s Hospital (IRB 00000277; Webb) as a reliant site to Yale. George Washington University (GWU) assumed the role of lead institution (IRB 031802; Pelphrey) and the ACE project is now governed as a single-site IRB at the University of Virginia (UVA; IRB-HSR-220423; Pelphrey). The ACE GENDAAR Data Coordination Center (DCC) was originally governed by UCLA, subsequently by the University of Southern California (USC; HS-18-00467, HS-13-00668; Van Horn), and presently by UVA (IRB-HSR-22078; Van Horn). All de-linked phenotypic, neuroimaging, EEG, and genetics data from ACE GENDAAR has been shared with the NIMH Data Archive (NDA), beginning 11/23/2012, under study collection IDs 2021 and 2804 where they are available for download and re-analysis by NDA-approved investigators. Written informed consent was obtained from parents and assent from children. General exclusion criteria included full-scale IQ (FSIQ) ≤70 (as estimated by the Differential Ability Scales–Second Edition General Conceptual Ability [DAS-GCA] score), twin status, active tic disorder interfering with imaging, pregnancy, presence of metal in the body, seizures within the past year, or current use of benzodiazepines, barbiturates, or anti-epileptic medications. Use of other medications was permitted only if dosage and regimen were stable for ≥6 weeks.

Additional exclusionary criteria specific to the non-autistic group were as follows: diagnosed, referred, or suspected ASD, schizophrenia, intellectual disability, learning disability, or other developmental or psychiatric disorder; a first- or second-degree relative with ASD; a total t-score >60 on the Social Responsiveness Scale, Second Edition^46^; a raw score >11 on the Lifetime version of the Social Communication Questionnaire^47^; clinical impression suggesting ASD, other developmental delay or disorder, broader autism phenotype, or significant psychiatric disorder.

Additional exclusionary criteria specific to the ASD group were as follows: known single gene disorder related to ASD or syndromic form of ASD (e.g., Fragile X); medical conditions likely to be etiological (e.g. focal epilepsy or infantile spasms); any neurological disorder involving pathology above the brainstem, other than uncomplicated non-focal epilepsy; history of significant pre- or perinatal injury, i.e. birth at <36 weeks and weight <2000 grams, or neonatal intensive care unit hospital stay >3 days; history of neonatal brain damage; any known environmental circumstances that might account for the picture of ASD in the proband (e.g., severe nutritional or psychological deprivation); clinically significant visual or auditory impairment after correction; or any sensorimotor difficulties that would preclude valid use of the diagnostic instruments.

Children and adolescents who received the Autism Diagnostic Observation Schedule, Second Edition (ADOS-2) Module 3 (for which revised algorithms were available from the outset of the study) were required to achieve a Calibrated Severity Score (CSS) of ≥4; adolescents who received Module 4 (for which a revised algorithm became available during the course of data collection^48^) were required to meet ASD criteria according to either the updated algorithm (CSS ≥4) or the version of the algorithm published with the ADOS-2 (Communication + Social Interaction Total ≥7). Scores on the Autism Diagnostic Interview-Revised (ADI-R), a standardized parent interview designed to obtain ASD symptom information, were required to meet the diagnostic algorithm within one point.

### 2.1. Sensory Profile

Sensory Profile self-report data were available from 223 autistic participants (ages 10-34 years) from two waves of data collection of an NIH-sponsored Autism Centers for Excellence research consortium. Data collections sites included five institutions: (1) the Center for Translational Developmental Neuroscience, Child Study Center, Yale School of Medicine, New Haven, CT; (2) the Nelson Laboratory of Cognitive Neuroscience, Boston Children’s Hospital, Harvard Medical School, Boston, MA; (3) the Center on Human Development & Disability, Seattle Children’s Hospital, University of Washington School of Medicine, Seattle, WA; (4) Staglin IMHRO Center for Cognitive Neuroscience, David Geffen School of Medicine, University of California, Los Angeles, CA; and (5) the Medical Imaging Research Center, Department of Radiology and Medical Imaging, University of Virginia, Charlottesville, VA. Both raw item-level responses (60 items) and four quadrant-level scores (low registration, sensation seeking, sensory sensitivity, sensation avoiding) were considered. Any missing data items were imputed using multivariate iterative imputation. Data were z-scored prior to analysis. Table 1 displays a full accounting of participant demographic information.

### 2.2. Functional Imaging

Blood-oxygen-level-dependent (BOLD) fMRI data were available from 113 autistic participants (ages 10-18 years) from four data collection sites using either Siemens 3T TrioTim or Siemens Prisma scanners equipped with 32-channel head-coils. Whole-brain T1-weighted MPRAGE images (FOV 256x256x256 mm³, 1x1x1 mm³ voxels, TE=3.3 ms, TR=2000 ms) and whole-brain gradient-echo EPI images (FOV 220x220x220 mm³, 3.4x3.4x4 mm³ voxels, TE=25 ms, TR=2000 ms) were collected. Preprocessing was conducted using fMRIPrep 23.0.2^49^ and included motion correction, slice-timing correction, skull-stripping, and spatial registration to MNI space. Denoising was performed using 3dTproject in AFNI^50,51^, and included the six motion realignment parameters and first six aCompCor^52^ components as nuisance regressors. Second-degree polynomial detrending and 0.01-0.1 Hz bandpass filtering were also performed.

To reduce the dimensionality of the fMRI data while focusing on a sensory-relevant region, only connections within and originating from the insula were considered (insula-seeded FC). Seed-based functional connectivity maps were computed for three insular subregions (dorsal anterior insula [dAI], ventral anterior insula [vAI], and posterior insula [PI]) for both resting state and the Biopoint task. For each sub-region, separate left and right seed maps were computed by correlating the seed time series with every brain voxel, applying Fisher *z*-transformation to correlation coefficients^53^, and then averaging the two maps. Connectivity maps were smoothed at 6 mm FWHM and analyzed within the MNI152 gray-matter mask^54^ thresholded at 0.30. These voxel-wise FC matrices were the inputs to the statistical models. Insula-seeded FC matrices were generated for both resting state and Biopoint tasks.

### 2.3. Statistical Methods: Subtyping and Spectral Embedding

Initially, both the item-level and quadrant-level data were subjected to two methods of subtyping: *k*-means and spectral subtyping (Ng-Jordan-Weiss algorithm^55^). The number of subtypes *k* was tuned from two to eight, and solutions were evaluated for silhouette score^56^, Calinski-Harabasz index^57^, Davies-Bouldin index^58^, and cross-validated adjusted Rand index^59,60^ (ARI) across 100 resamples; the top-performing *k* was determined by silhouette score. Both linear (PCA) and nonlinear (spectral) decompositions were used to assess the intrinsic dimensionality of the sensory space. PCA was used to assess linear variance structure, spectral embedding was used to intrinsic manifold topology under a graph-based similarity structure. The PCA scree plots demonstrate the dominant dimensionality of the data under linear assumptions. For diagnostics, a *k*-nearest-neighbor graph (*k*=10) was constructed on the standardized sensory features, and the eigenvalues of the normalized graph Laplacian were computed. The resulting spectral scree plots visualize the smallest Laplacian eigenvalues, where smooth eigenvalue growth indicates continuous manifold structure and large gaps would suggest discrete subtypes. To assess the robustness of the spectral embedding to graph construction, we evaluated the stability of the first spectral eigenvector across *k*-nearest-neighbor values ranging from 5 to 15. Feature importance was measured using Spearman’s rank correlation coefficient^61^ (*ρ*) to quantify the degree of monotonic alignment between an observed feature and the dominant latent sensory axis. The extracted spectral embeddings were then used as continuous sensory scores for downstream neuroimaging analyses. Item-level data were evaluated for feature importance to establish which areas of sensory experience most contributed to differentiation along the sensory dimension.

### 2.4. Statistical Methods: Voxel-wise fMRI Analysis

Analyses were restricted to autistic participants (63 total, 32 females, 50.79%). For each participant with multiple scans, we selected a single Biopoint per participant using a tiered quality-control procedure prioritizing runs that met volume threshold criteria and exhibited the lowest mean framewise displacement (FD). Participants with missing FD, sensory data, or covariates were excluded. The primary predictor of interest for each seed and fMRI task was the first sensory spectral eigenvector, derived from item-level spectral embedding of Sensory Profile responses. This predictor was treated as a continuous measure indexing individual differences along the dominant sensory regulation axis.

The first spectral eigenvector was used as the primary sensory predictor because it accounted for the dominant dimension of variation in the item-level sensory data (see Figure 1) and demonstrated robust stability across *k*-nearest-neighbor graph constructions (Table 3). Higher-order eigenvectors were examined for completeness but did not exhibit comparable dominance in the Laplacian spectrum and were therefore not included as primary predictors in the voxel-wise models.

**Figure 1:**
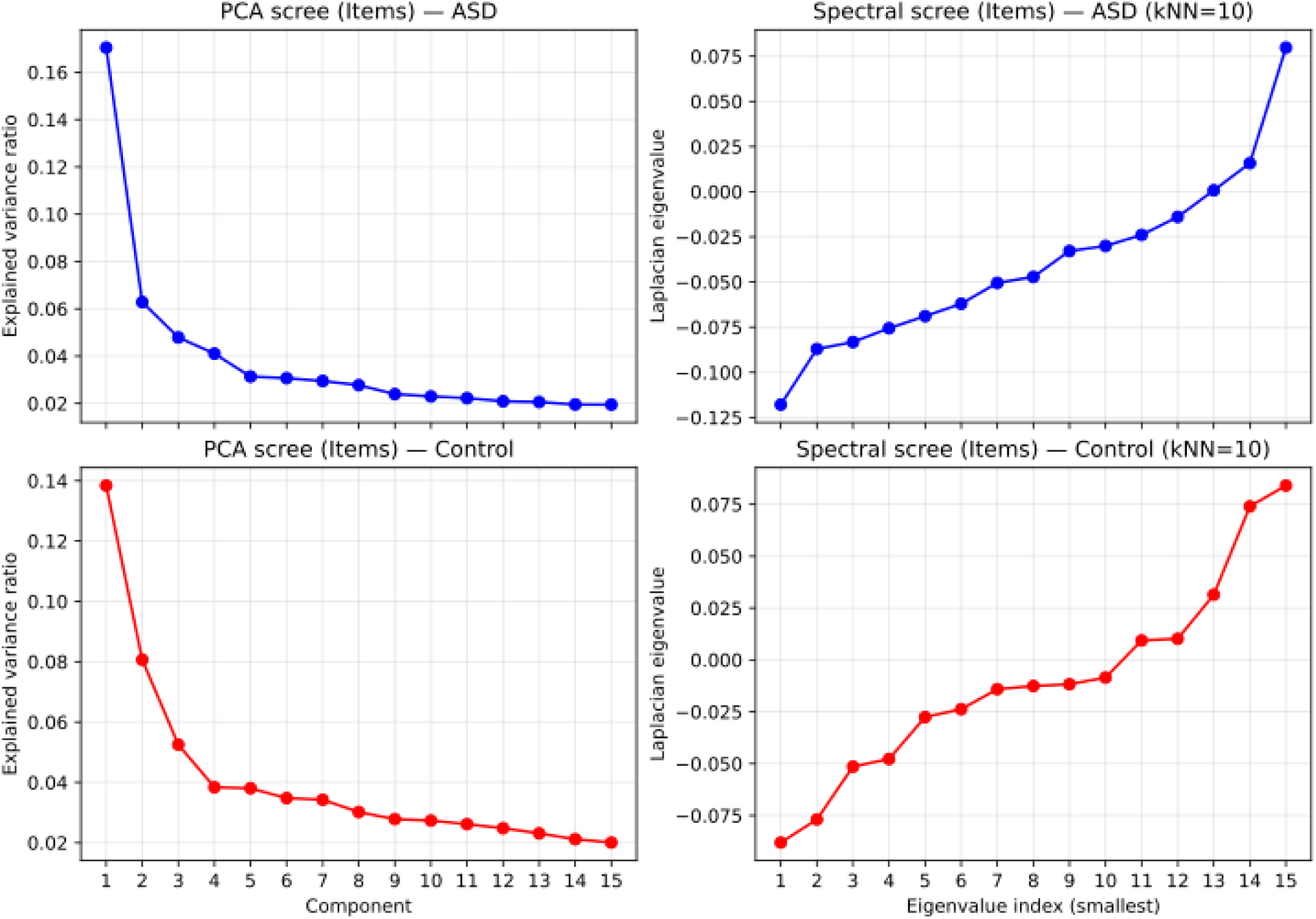
Linear (PCA) scree plots (left) and nonlinear (spectral) scree plots (right) for the item-level sensory data. Autistic participant plots are presented on the top row in blue, non-autistic participants on the bottom row in red. Unidimensional structure is confirmed in these plots due to the dominance of the first PCA eigenvalue as well as the smooth initial increase in Laplacian eigenvalues.

Voxel-wise linear mixed-effects models were fitted separately for each insula seed using AFNI’s 3dLMEr^50,51,62^. The model included fixed effects for the sensory spectral eigenvector, sex, and their interaction, along with covariates for scan age, IQ (measured using Differential Abilities Scale – Greater Conceptual Ability score [DAS-GCA]), age-residualized Pubertal Development Scale total score (PDS), mean framewise displacement, and data collection site. Collection site was modeled as a random effect to account for unknown scanner effects; two sites upgraded scanners halfway through the study, leading them to be considered as separate sites pre- and post-upgrade and thereby imposing no assumptions about harmonization. Although functional imaging data were available for 113 autistic participants, voxel-wise analyses were restricted to participants with complete sensory, motion, and covariate data.

Because whole-run FC estimates computed during task paradigms can be artificially inflated by shared task timing or block-locked coactivation, a targeted sensitivity analysis was conducted to ensure our insula-seeded FC results reflected intrinsic coupling rather than unmodeled task structure. To test this, we randomly selected a sub-sample of participants (*n*=10) from the final analysis cohort and re-computed their FC maps after explicitly regressing out the task timing.

For each of these participants, exact task timing for the Biopoint paradigm was reconstructed from raw ePrime logs to generate TR-locked boxcar regressors corresponding to the two task conditions (biological motion; random motion). These unconvolved task regressors were added to the original nuisance regression model (which included motion parameters, their temporal derivatives, anatomical CompCor components, and polynomial drift terms) during the voxel-wise residualization of the preprocessed fMRI data. Whole-run voxel-wise FC maps were then recomputed for the PI, dAI, and vAI seeds, yielding paired FC maps for each participant and seed: one with standard nuisance regression, and one with standard nuisance plus explicit task regression.

## 3. Results

### 3.1. Sensory Subtyping

Across each subtyping methodology (*k*-means, spectral), feature-level inputs (items, quadrants), and diagnostic condition (autistic, non-autistic), the results suggest weak separation and limited stability. For *k*-means, silhouette scores routinely displayed poor separability (silhouette scores≤0.10 in ASD; ≤0.08 in non-ASD). Meanwhile, the spectral subtyping tended toward modest stability (ARI≤0.34 in both diagnostic conditions) without improvement in terms of separability. The choice of *k* was also inconsistent, varying mostly by subtyping method. These results suggest the data were quite sensitive to methodology and did not produce evidence of a salient subtype structure. Table 2 shows the full results for each subtyping attempt and each value of *k* tested.

Both linear (PCA) and nonlinear (spectral) decompositions, displayed as scree plots in Figure 1, reveal a dominant first dimension in the Sensory Profile data. The results of the spectral decomposition were robust to perturbation resampling and across tested *k*-nearest-neighbor settings (Table 3). This interpretation is illustrated in Figure 2 where the same data that produce unstable subtypes organize into a continuous manifold when visualized with overlaid sensory quadrant scores. The top plots (autistic participant data) demonstrate the unidimensional nature of the sensory data by showing a smooth increase along the horizontal axis for quadrants 1, 3, and 4. The bottom plots (non-autistic participant data) do not follow this same pattern, thus revealing an ASD-specific sensory response. Interestingly, quadrant 2 appears to show a unidimensional structure along the second spectral dimension in the non-ASD data that is not present in the ASD data.

**Figure 2:**
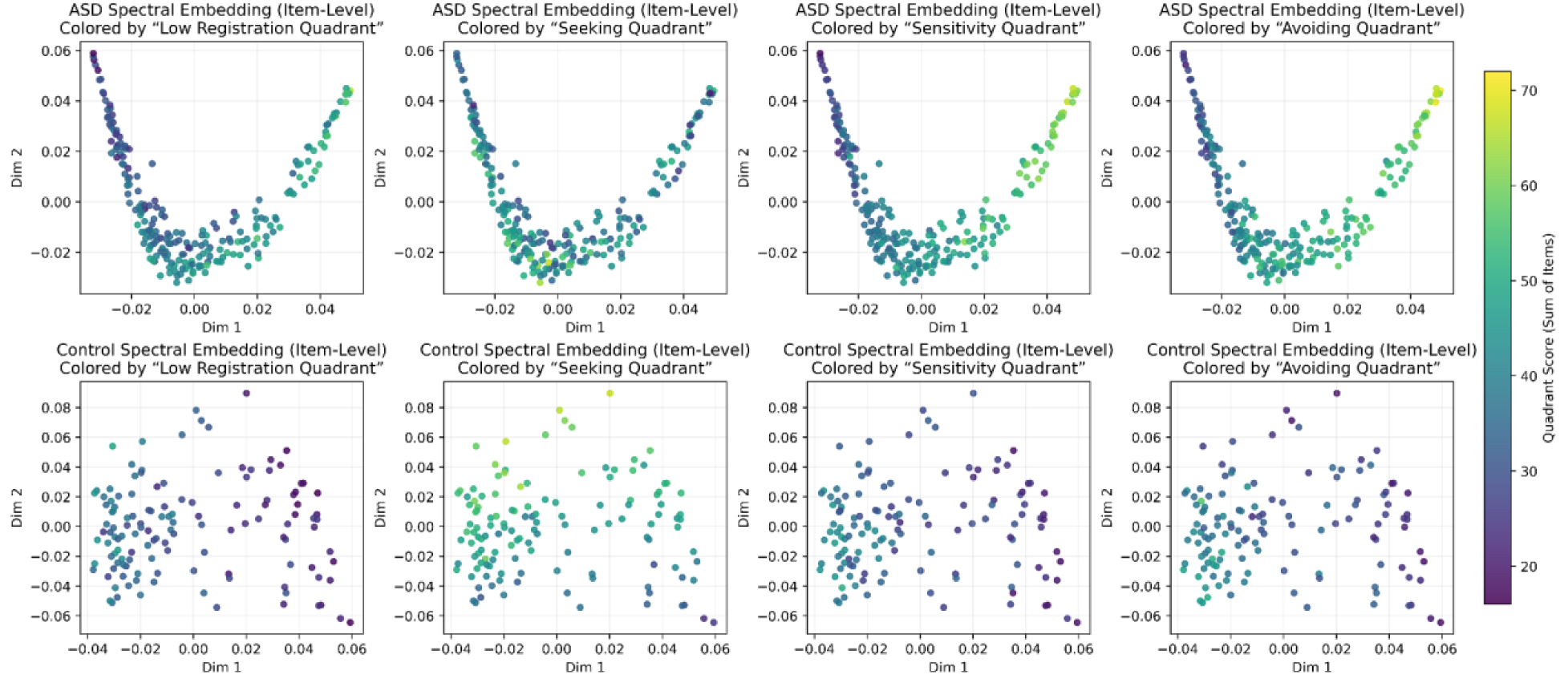
Spectrally embedded scatterplots colored by quadrant score. Top: autistic participants; bottom: non-autistic participants.

Table 4 displays the Spearman rank correlations between the first sensory dimension and demographic and behavioral measures within the imaging ASD cohort (*n*=63). Age was the only variable significantly associated with the sensory dimension (*ρ*=0.380; *p*=0.003), indicating modest developmental variation along the sensory axis. No significant associations were observed for biological sex, IQ (DAS-GCA), ADOS Calibrated Severity Score (CSS), Repetitive Behavior Scales Revised (RBS-R) total score, Child Behavior Checklist (CBCL) total score, Behavior Rating Inventory of Executive Function Global Executive Composite (BRIEF-GEC), and the Social Responsiveness Scale (SRS) total score (*p*>0.08 for all)^46,63–65^. We also performed partial correlational (partial Spearman correlations controlling for age, sex, IQ, and ADOS CSS) analyses to examine whether the sensory dimension reflected symptom severity or general cognitive factors broadly. These analyses likewise revealed no significant associations between the sensory dimension and any behavioral score (*p*>0.58 for all).

Feature importance, quantified using Spearman’s *ρ* on the first spectral dimension, showed that the strongest contributors to the sensory spectral score came primarily from the Sensory Sensitivity and Sensory Avoiding quadrants. The top 15 items in the ASD group accounted for 43.9% of the overall loading mass; the top 15 items in the non-ASD group accounted for 39.1% of the overall loading mass. A full accounting of the feature importance scores for all 60 Sensory Profile items can be found in Tables 5 (ASD) and 6 (non-ASD).

In ASD, the features most strongly aligned with the primary spectral axis reflect anticipatory sensory regulation, including avoidance of crowds and noisy environments, distress in visually dense contexts, and proactive strategies to dampen sensory input. In contrast, control feature alignments were dominated by context-specific processing difficulties (e.g., following rapid speech, sustaining attention), without convergence onto a common regulatory theme.

Sensory severity distributions were comparable between the full behavioral cohort (*n*=223) and the imaging subsample (*n*=63) for both sexes (Figure 9 in Appendix). The imaging cohort spanned the central range of the full distribution without evidence of truncation or sex-specific skew, indicating that subsequent neuroimaging analyses were not driven by range restriction or sampling bias.

### 3.2. Functional Imaging

Linear mixed-effects modeling revealed clusters of significant interactions between the sensory spectral score and sex in both the dAI seed (*q*=0.009) and the PI seed (*q*=0.049) connectivity maps during the Biopoint task, with no such effects observed in the resting state task (*q*≈1.0 for both). In the vAI seed, biological sex emerged as the only significant predictor in both scan conditions (Biopoint *q*=0.004; resting state *q*=0.020).

To evaluate whether higher-order sensory dimensions contributed additional explanatory value, voxel-wise models were also estimated using the second spectral eigenvector. No significant main or interaction effects survived correction for multiple comparisons for any seed or scan condition, and inclusion of the second eigenvector did not alter the pattern of effects observed for the first dimension. These findings support the interpretation that the observed connectivity associations are driven primarily by the dominant sensory regulation axis.

To control for false positives while preserving sensitivity to spatially extended effects, cluster-level thresholding was applied, yielding a final set of statistically significant interaction clusters for each seed. Voxel-wise statistical maps corresponding to the interaction term were established using a cluster-forming threshold of *p*<0.005 (two-tailed, minimum cluster size=20 voxels).

For each of the 13 clusters (6 from dAI, 7 from PI), cluster size (in voxels), peak voxel coordinates, maximum *z*-statistic, and center-of-mass coordinates were extracted (Table 7). Macro-anatomical labels were identified using AFNI’s “whereami” tool with the Eickhoff-Zilles N27 macro labels atlas^66–68^. Clusters were identified bilaterally in postcentral, precentral, frontal, temporal and occipital gyri as well as in the cerebellum. Subject-level mean connectivity values were extracted from each significant cluster by computing the mean Fisher-*z*-transformed value across all voxels in that cluster. Axial, coronal, and sagittal montage views of these clusters are presented in Figures 3 and 4.

**Figure 3:**
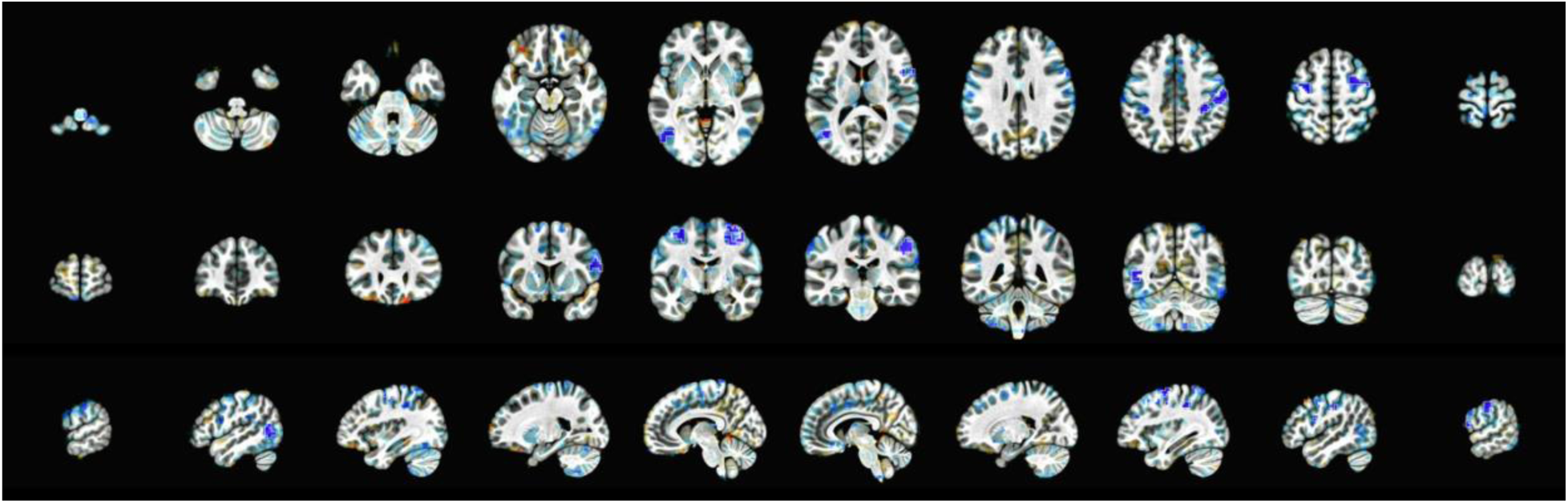
Axial (top), coronal (middle), and sagittal (bottom) views of the significant sensory gradient and sex interaction clusters in the dAI seed. Blue coloration indicates areas where the relationship (slope) between dAI connectivity and the sensory gradient was significantly more negative in autistic males than in autistic females; red coloration indicates the opposite effect.

**Figure 4:**
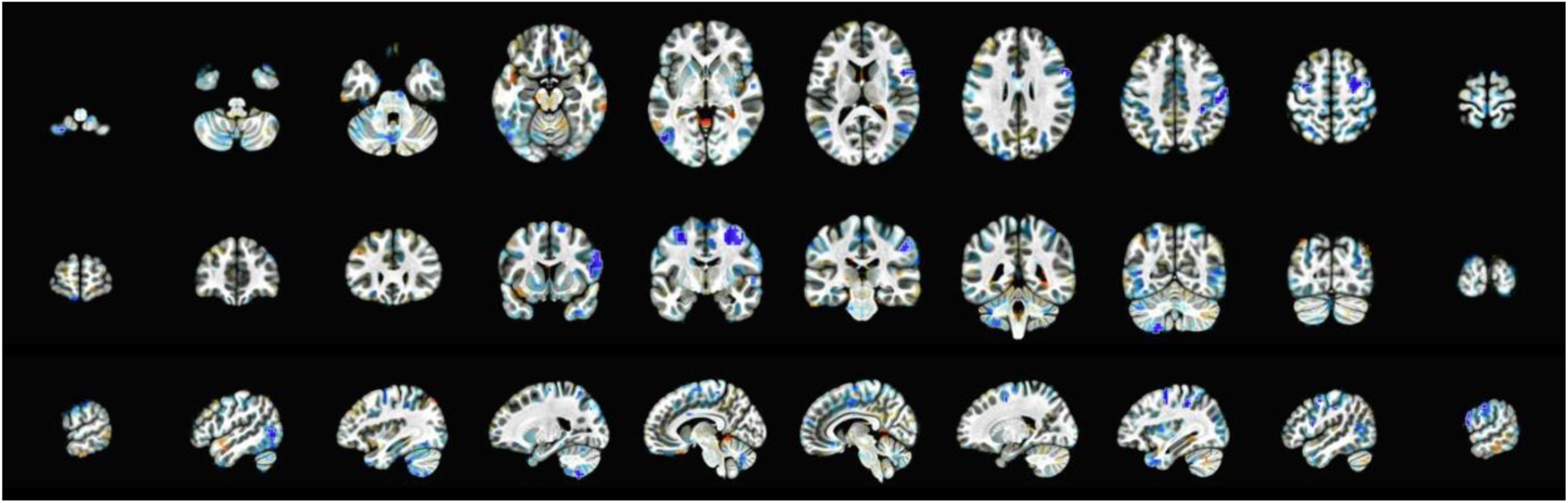
Axial (top), coronal (middle), and sagittal (bottom) views of the significant sensory gradient and sex interaction clusters in the PI seed. Blue coloration indicates areas where the relationship (slope) between PI connectivity and the sensory gradient was significantly more negative in autistic males than in autistic females; red coloration indicates the opposite effect.

The voxel-wise mixed-effects modeling analysis established significant interaction effects in the dAI and PI seeds but did not explicitly indicate directionality. To investigate this, post-hoc visualizations of the sensory spectral score per cluster, stratified by sex, were created (Figures 5 and 6; Appendix 1). We visualized cluster-averaged connectivity values as a function of sensory severity, stratified by sex. In both dorsal anterior and posterior insula seeds, increasing sensory severity was associated with reduced insula–motor cortex connectivity in both sexes; however, this relationship was markedly steeper in males than females, consistent with the direction of the voxel-wise interaction effects.

**Figure 5:**
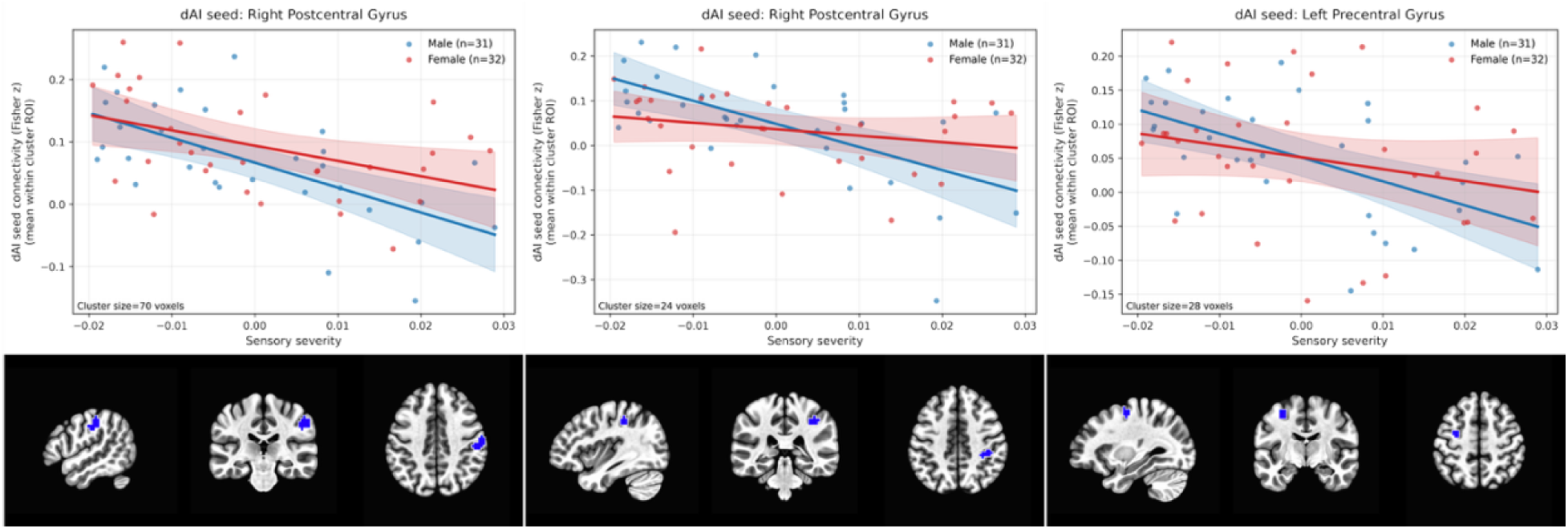
Scatterplots showing the relationship between sensory severity (first spectral eigenvector) and insula-seeded functional connectivity for dAI during the biological motion task. Each point represents an individual participant, stratified by sex (blue=male, red=female). Solid lines indicate sex-specific linear fits with shaded 95% confidence intervals. Brain slices depict the anatomical location of the cluster. The left and middle panels represent the somatosensory cortex (cluster sizes=70 and 24 respectively); the right panel represents the primary motor cortex (cluster size=28). In each of these regions, the primary effect of sensory gradient was significant in addition to the sensory severity-by-sex interaction.

**Figure 6:**
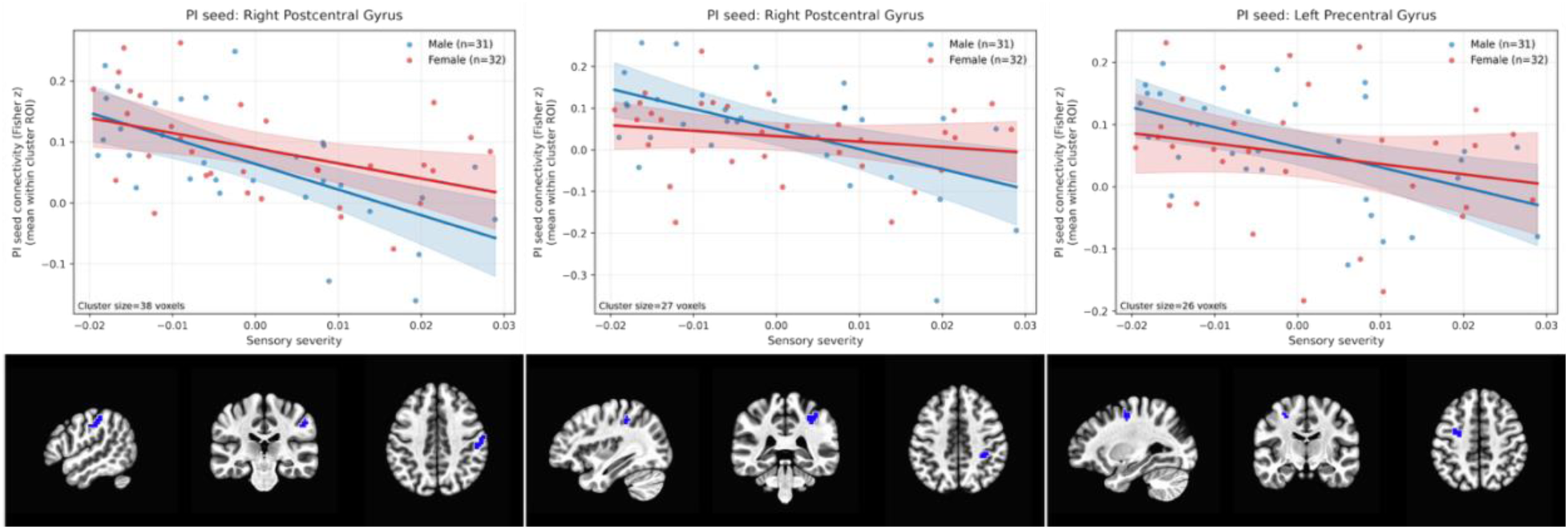
Scatterplots showing the relationship between sensory severity (first spectral eigenvector) and insula-seeded functional connectivity for PI during the biological motion task. Each point represents an individual participant, stratified by sex (blue=male, red=female). Solid lines indicate sex-specific linear fits with shaded 95% confidence intervals. Brain slices depict the anatomical location of the cluster. The left and middle panels represent the somatosensory cortex (cluster sizes=38 and 27 respectively); the right panel represents the primary motor cortex (cluster size=26). These findings are confirmatory of those in the dAI seed.

To evaluate whether the observed sex-by-sensory interaction effects were driven by individual leverage points, we conducted influence diagnostics and leave-one-out (LOO) resampling analyses on subject-level cluster mean connectivity values for all significant clusters. The primary sensorimotor clusters reported in the main text (dAI clusters 2, 5, 6; PI clusters 3, 4, 5) exhibited stable interaction coefficients under LOO resampling. Across these clusters, the interaction term remained negative in all 63 LOO iterations, indicating that no single participant reversed the direction of the effect (Table 8).

In contrast, several smaller or peripheral clusters demonstrated greater variability in interaction estimates, including occasional sign reversals under LOO resampling. These clusters were not emphasized in primary interpretations. Maximum Cook’s D values ranged from 0.13 to 0.45 across clusters, with 0–6 subjects exceeding the heuristic (4/*n* = 0.0635) threshold. However, in all primary clusters, removal of high-leverage observations did not alter the direction of the interaction term. This is reported in full in Table 9.

To ensure the integrity of the functional connectivity results, we performed a rigorous assessment of motion and signal-to-noise ratio (SNR) within the imaging cohort. We compared head motion (mean framewise displacement) and temporal SNR (tSNR) across states and evaluated their relationship with our primary variable of interest, the sensory dimension. Significant state-level differences were observed; specifically, resting state showed a higher mean FD and higher tSNR compared to the biological motion state (*p*=0.010; *p*<0.001 respectively). However, neither mean FD nor tSNR correlated with sensory severity in either the biological motion or resting states (all |*ρ*|≤0.07; *p*≥0.62; see Table 10). To further mitigate the impact of transient motion artifacts, all primary connectivity analyses utilized mixed linear models with mean FD included as a nuisance covariate. Taken together, these results demonstrate that the observed relationships between sensory severity and functional connectivity are not driven by spurious motion artifacts or global differences in signal quality.

Finally, to quantify the impact of task regression, we computed the voxel-wise spatial similarity (Pearson *r*) and the mean difference in Fisher-*z* values (ΔZ) between the standard and task-regressed FC maps within the whole-brain mask. Across all participants and insular subregions, task regression had a negligible effect on FC topography, demonstrated by the high spatial correlation between the standard and task-regressed maps (mean Pearson *r*=0.991, minimum=0.973). Furthermore, the magnitude of connectivity was quite small, with the mean voxel-wise change in Fisher-*z* values approaching zero (mean ΔZ=0.0006±0.016). Because modeling the task structure did not meaningfully alter the FC estimates, all primary analyses were conducted using the standard nuisance-only model to preserve temporal degrees of freedom.

## 4. Discussion

### 4.1. Sensory Variation as a Continuous Regulatory Manifold

The present findings indicate sensory sensitivity in ASD follows a continuous latent dimension and does not separate into distinct sensory subtypes. The failure of subtyping is therefore informative rather than inconclusive. The sensory spectral score, or sensory gradient, captures a coherent pattern of over-reactivity and avoidance that aligns with well-established autistic behavioral traits like sensory overload and inability to handle unpredictable environments^69^. Autistic individuals may attempt to regulate sensory inputs by anticipating and avoiding such environments, many of which involve social proximity. This indicates that categorical phenotyping imposes artificial boundaries on behavior that is inherently continuous and nonlinear.

Our results indicate that sensory traits are constrained by a common regulatory strategy in ASD, whereas in non-autistic individuals, they vary independently across contexts. The feature importances in conjunction with the spectral scatter plots reveal that autistic sensory traits are organized along a gradient aligned with sensory sensitivity and sensation avoidance. The same cannot be said of the non-autistic participants, whose spectral embeddings do not reveal a clear manifold structure, feature importances tend to be more context-specific, and they do not adhere to a common low-dimensional structure.

Although PCA also demonstrated a dominant first component, the crescent-shaped geometry of the ASD spectral space (Figure 2) reflects a nonlinear, low-dimensional sensory regulation manifold not captured by linear projections. Increasing sensory sensitivity results in compensatory avoidance behaviors, whereas non-autistic participants do not exhibit a comparable constrained structure. In fact, the manifold structure for the non-autistic participants suggests their sensory experience is multidimensional, evidenced by the smooth gradient along the second latent dimension (y-axis) in the Sensory Seeking quadrant (visualized in Figure 2). This may be indicative of a distinct sensory motivator, whereas in the autistic participants, this is collapsed into the general severity manifold.

### 4.2. Context-Dependent Integration

Most importantly, this severity gradient revealed specific neurobiological relevance during the perception of biological motion. The alignment between the item-level feature loadings, characterized by socially dense and unpredictable environments, and the stimuli-specific fMRI results suggests that sensory severity in ASD is linked to social context. If ASD sensory differences were global gain abnormalities, effects should appear at rest. Instead, sensory severity predicts connectivity only during socially embedded sensory processing, consistent with the existing literature^70,71^. The absence of significant connectivity associations at rest and their robustness during biological motion perception imply that the sensory gradient reflects a disruption in the integration of social stimuli rather than an intrinsic deficit. Together, the behavioral and neurological findings converge on a model in which sensory severity reflects difficulty integrating socially embedded sensory information.

### 4.3. Insula-Sensorimotor Hypoconnectivity

The observed hypoconnectivity between the insula and the primary motor and somatosensory cortices may represent a compensatory decoupling. Individuals with high sensory sensitivity appear to experience salience overload in social contexts; the resulting hypoconnectivity in insula-motor coupling could represent impacted integration between interoceptive processing or salience detection (dorsal anterior insula responsibilities)^72^ and motor simulation systems (primary motor cortex)^73^ when viewing biological motion. Such hypoconnectivity could influence the ability to translate sensory-salience input into coordinated motor readiness states. Biological motion is a stimulus that demands such integration, so the observed hypoconnectivity suggests a failure or decoupling of this pathway under high sensory load. Importantly, this pattern does not indicate motor impairment, but impaired translation of salience into action preparation under social sensory conditions.

### 4.4. Sex-Wise Moderation between Sensory Severity and Insula Hypoconnectivity

The significant interaction between biological sex and the sensory gradient suggests that the association between sensory severity and insula hypoconnectivity is significantly more negative in autistic males than in autistic females. Autistic females showed a weaker relationship between sensory scores and insula-motor coupling, potentially suggesting either a different neural pathway for sensory regulation, a greater degree of resilience to sensory overload, or strengthened ability to compensate for hypoconnectivity along this pathway. Given that autistic females exhibit the same profile of sensory severity as autistic males, this finding suggests that reported social “masking” and symptom camouflaging by autistic females may have a neurobiological component, consistent with proposed female protective effect and camouflaging mechanisms in ASD^74,75^.

The stability analyses further suggest that the observed sex-by-sensory interaction is concentrated within core sensorimotor regions rather than diffusely distributed across all 13 significant clusters. Primary motor and somatosensory clusters exhibited consistent interaction direction under leave-one-out resampling, whereas smaller peripheral clusters showed greater coefficient variability. This pattern is consistent with the hypothesis that sensory severity modulates insula–sensorimotor coupling in a circuit-specific manner rather than reflecting a global connectivity perturbation.

### 4.5. Implications for Sensory Phenotyping and Clinical Neuroimaging

The transition from categorical subtyping to dimensional gradients has profound implications for the Research Domain Criteria framework in psychiatry^76,77^. The present results demonstrate that traditional person-centered modeling (like *k*-means) can result in misleading interpretations if the underlying data topology is better represented as a continuous manifold. In clinical trials, subtyping is often used to select participants for interventions; however, if these subtypes are unstable or artifacts of the selected subtyping algorithm, the trial cannot be considered valid or actionable.

From a clinical neuroimaging perspective, this study suggests that “sensory features” should not be treated as a static covariate. Instead, neuroimaging protocols should prioritize stimulus-evoked designs that specifically interrogate the integration of salience and action. Otherwise, a major phenotypic driver of sensory disability in ASD can be overlooked.

### 4.6. Limitations and Future Directions

Together, these results suggest that sensory severity in ASD reflects a constrained regulatory manifold whose neural consequences emerge selectively under socially demanding conditions and are differentially modulated by sex. Some future directions include explorations of sensory-based functional connectivity differences in other task-based scans. For instance, in this project, autistic participants completed five functional tasks; the other three (face identification, language, and social reward) could interact with sensory severity in different ways.

Furthermore, the sensory severity gradient should be quantified and validated by association with other behavioral outcome measures, including CBCL or BRIEF subscales. The sensory severity gradient was not significantly associated with the total scores for these measures, but the granularity of the subscales could uncover meaningful associations. Our previous work has found a number of correlations between the raw scores of these subscales, and future efforts should be directed at uncovering gradients and latent factors underlying larger behavioral measures^39^. Further confirmation of this effect and its specificity to ASD should be explored as well. While non-autistic participants were analyzed behaviorally to establish the uniqueness of the ASD manifold, a critical next step is to run comparative voxel-wise LME models on the control group. The expected results would be a distinct, unconstrained sensory severity manifold as well as no significant relationship in the sensorimotor brain regions.

A primary limitation is the Sensory Profile’s inherent design, which conflates sensory thresholds with behavioral regulation. While the spectral embedding successfully mapped a regulatory manifold, future work should utilize instruments that separate purely perceptual thresholds from downstream affective responses to examine whether this manifold structure persists at early stages of sensory encoding.

Another limitation of this analysis is that the insula-seeded regions were averaged laterally, potentially limiting the granular detail that could emerge from splitting the seeds laterally. The decision to average the left and right hemispheres was made to ease the computational and multiple comparison burdens, but the strong interaction effect is encouraging and warrants more fine-grained lateral exploration.

### 4.7. Conclusion

In conclusion, this study provides a novel framework for parsing the complex heterogeneity of sensory processing in ASD. These findings demonstrate that the failure to identify reproducible sensory subtypes in the literature may be a consequence of categorical modeling rather than biological reality. By treating sensory responsivity as a latent manifold, we identified a robust behavioral-to-neural connection that traditional subtyping methods obscured. Context-dependent decoupling within the insula-motor loop emerged during the integration of socially relevant sensory information, underscoring the critical importance of aligning behavioral modeling with the intrinsic topology of clinical data. This study serves as a proof-of-concept for the necessity of hypothesis-driven data science in clinical neuroimaging, illustrating that the discovery of neural biomarkers is fundamentally dependent on the mathematical precision of behavioral phenotyping.

## Data Availability

All data used in this study are publicly available at the NIMH Data Archive (NDA) under the collections #2021 and #2804.

https://nda.nih.gov/edit_collection.html?id=2021

https://nda.nih.gov/edit_collection.html?id=2804

## Appendix: Tables and Figures

**Figure 7:**
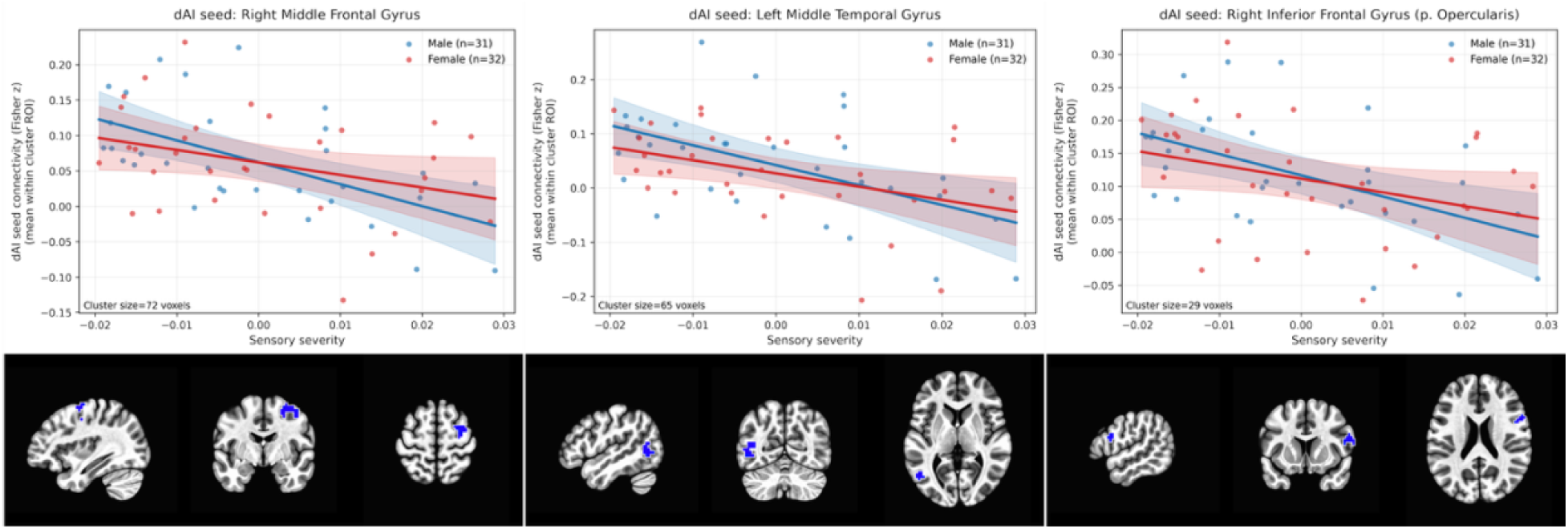
Scatterplots showing the relationship between sensory severity (first spectral eigenvector) and insula-seeded functional connectivity for dAI during the biological motion task. Each point represents an individual participant, stratified by sex (blue=male, red=female). Solid lines indicate sex-specific linear fits with shaded 95% confidence intervals. Brain slices depict the anatomical location of the cluster. Left: dAI cluster 1, representing the right middle frontal gyrus (cluster size=72 voxels). Middle: dAI cluster 3, representing the left middle temporal gyrus (cluster size=65 voxels). Right: dAI cluster 4, representing the right inferior frontal gyrus (p. opercularis; cluster size=29 voxels).

**Figure 8:**
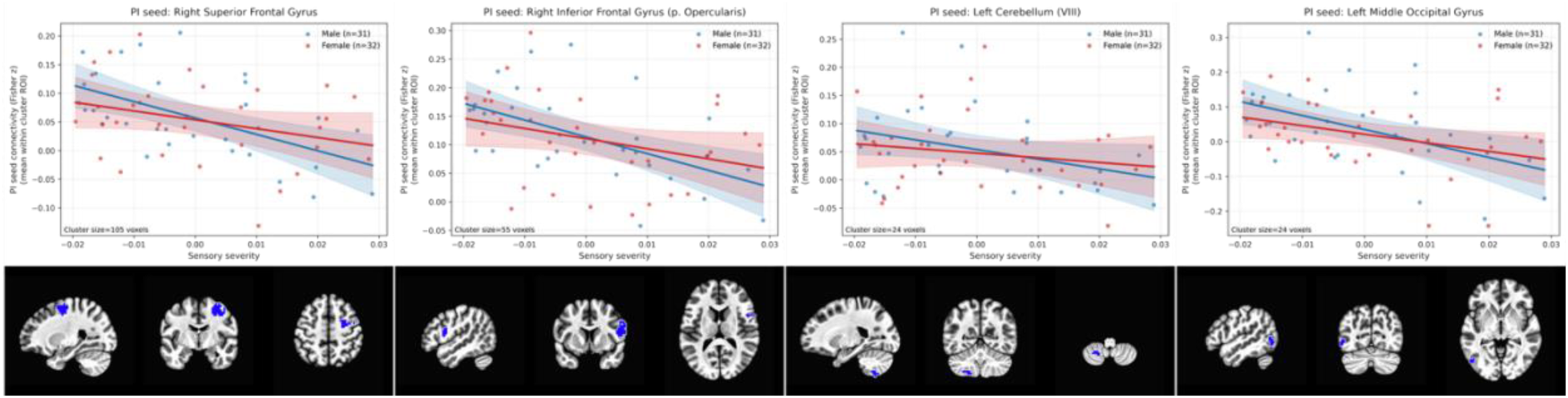
Scatterplots showing the relationship between sensory severity (first spectral eigenvector) and insula-seeded functional connectivity for PI during the biological motion task. Each point represents an individual participant, stratified by sex (blue=male, red=female). Solid lines indicate sex-specific linear fits with shaded 95% confidence intervals. Brain slices depict the anatomical location of the cluster. Left: PI cluster 1, representing the right superior frontal gyrus (cluster size=105 voxels). Middle left: PI cluster 2, representing the right inferior frontal gyrus (p. opercularis; cluster size=55 voxels). Middle right: PI cluster 6, representing the left cerebellum (cluster size=24 voxels). Right: PI cluster 7, representing the left middle occipital gyrus (cluster size=24 voxels).

**Table 1:**
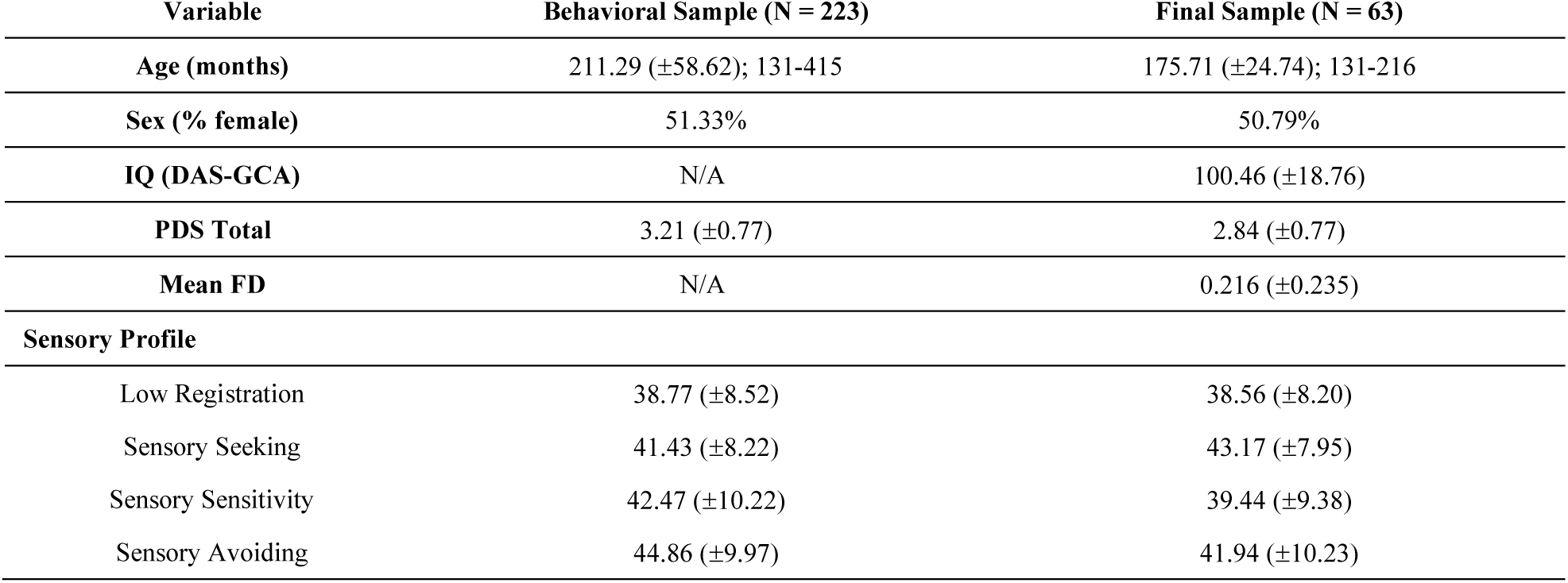
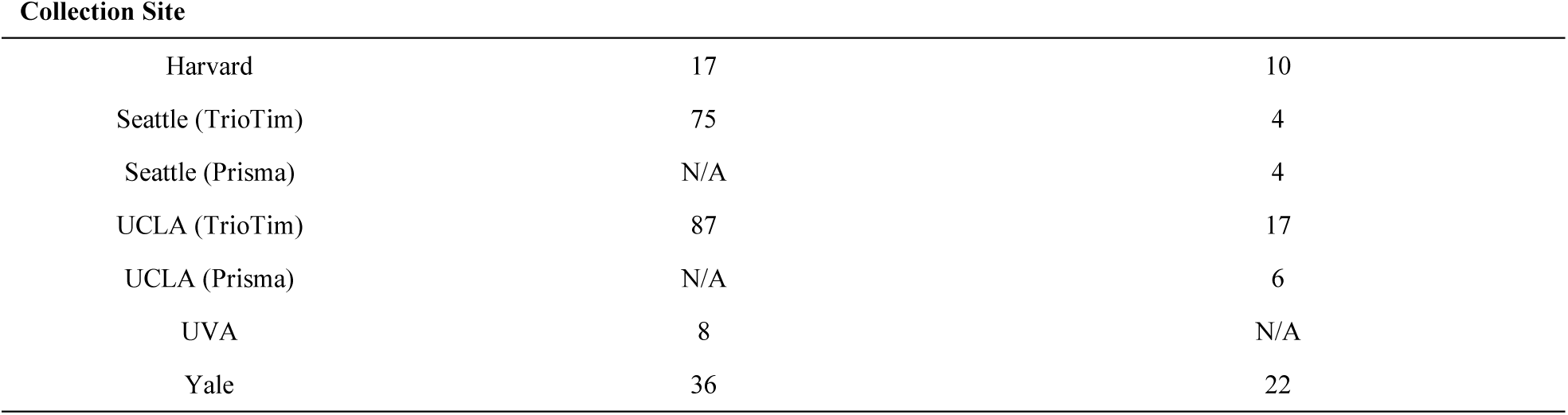
Demographic characteristics of the dataset. IQ is not reported for the behavioral sample due to inconsistent IQ measurement across Waves 1 and 2 of data collection. Mean FD is also not reported for the behavioral sample due to irrelevance to the data analyzed. UVA was not <u>involved in the data collection of Wave 1 and thus did not have any samples present in the final cohort.</u>

**Table 2:**
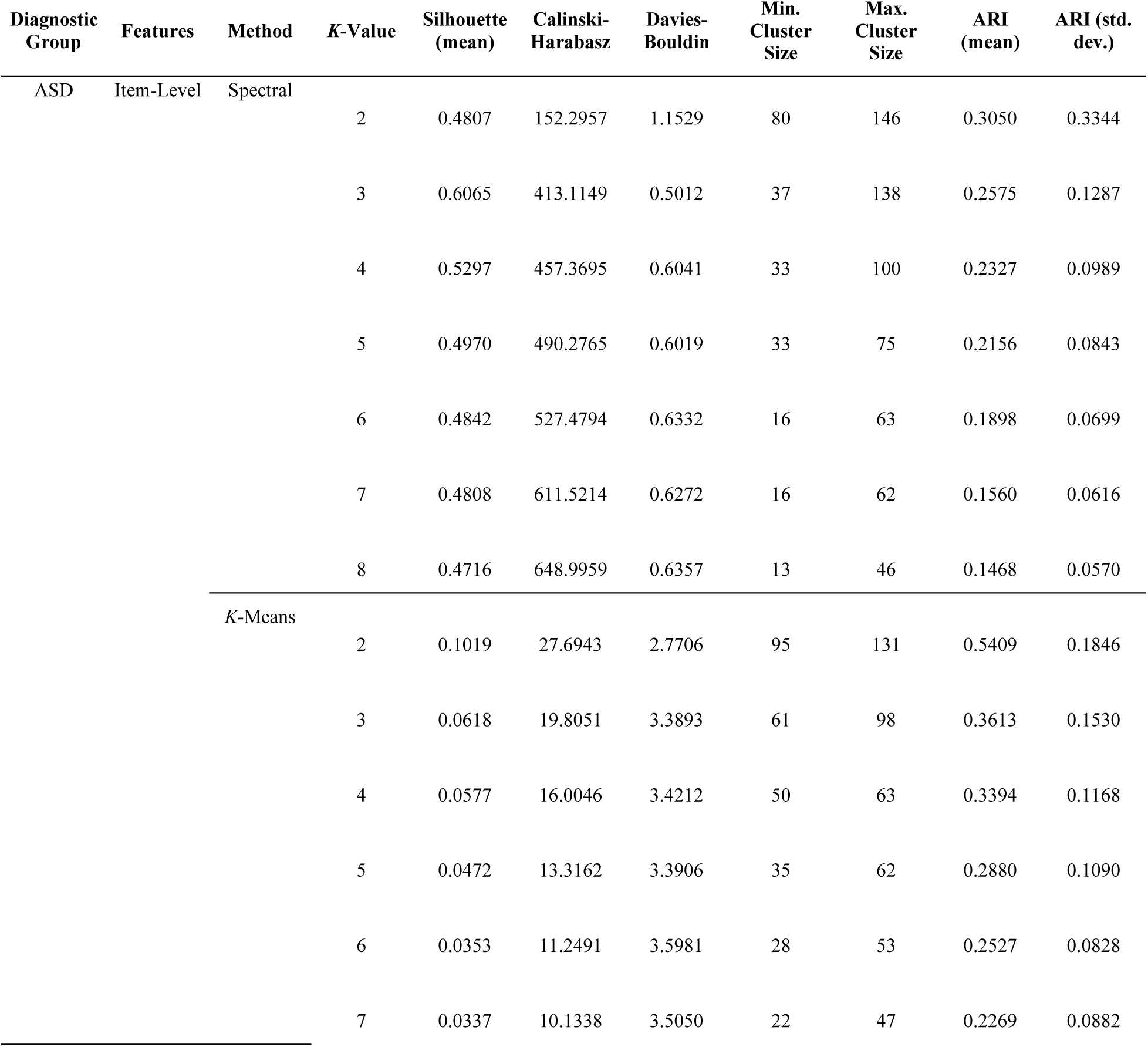
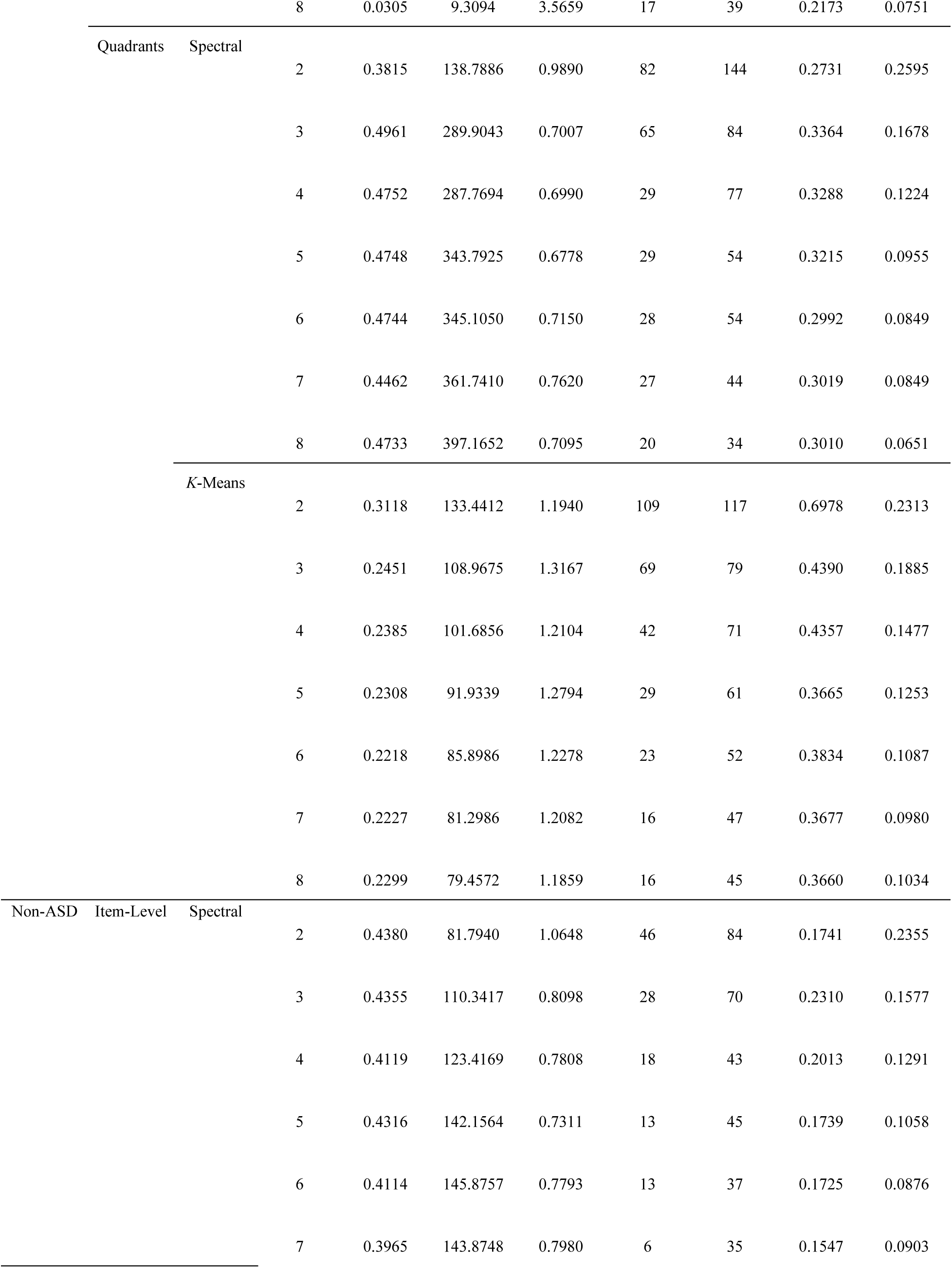
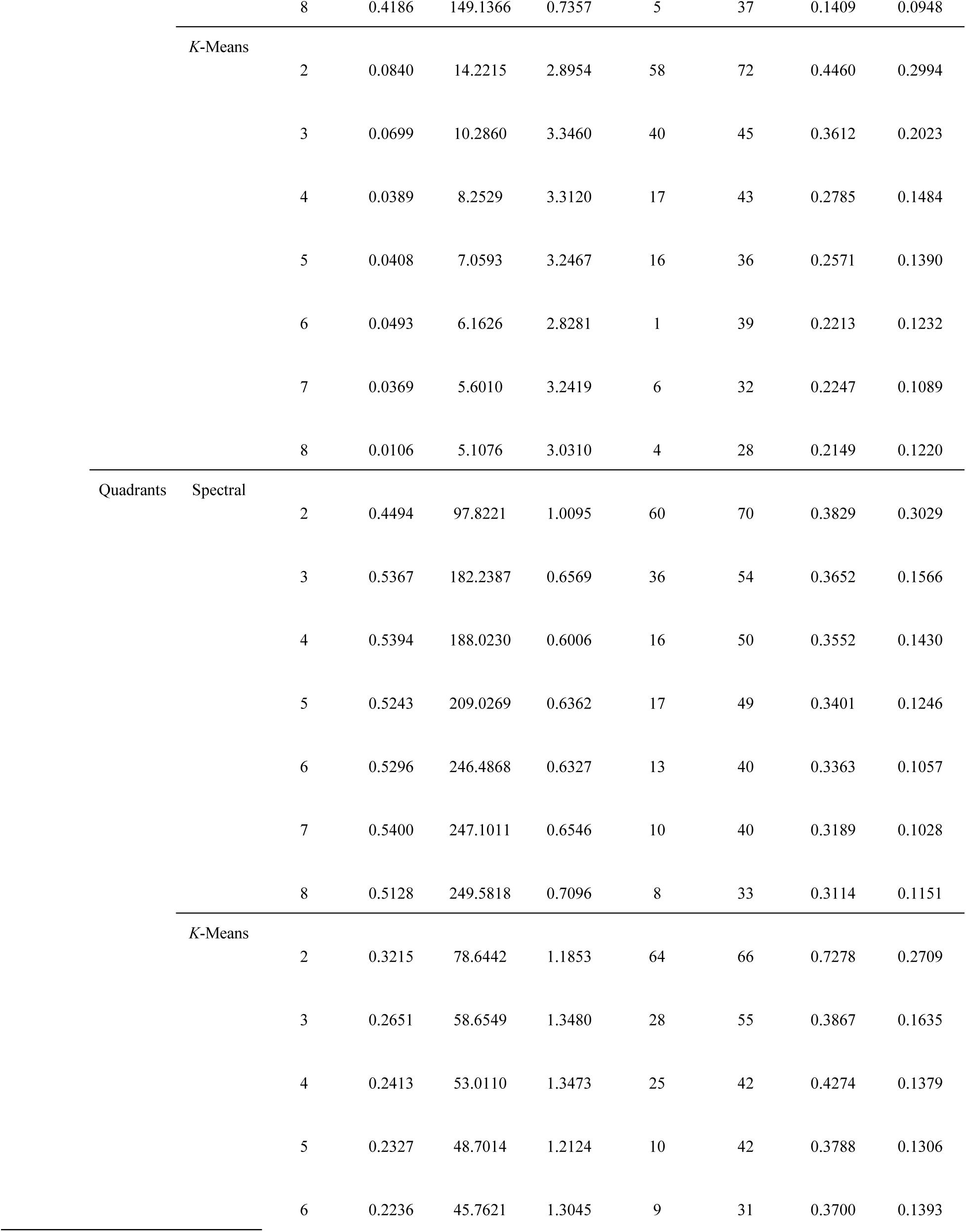
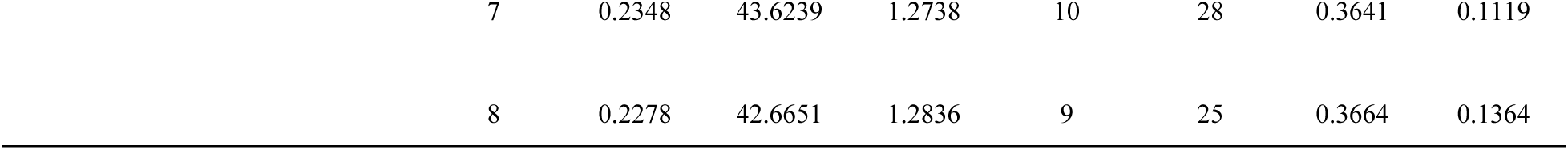
Full list of results from the subtyping methods, organized by diagnostic group, then feature level, then algorithm.

**Table 3:**
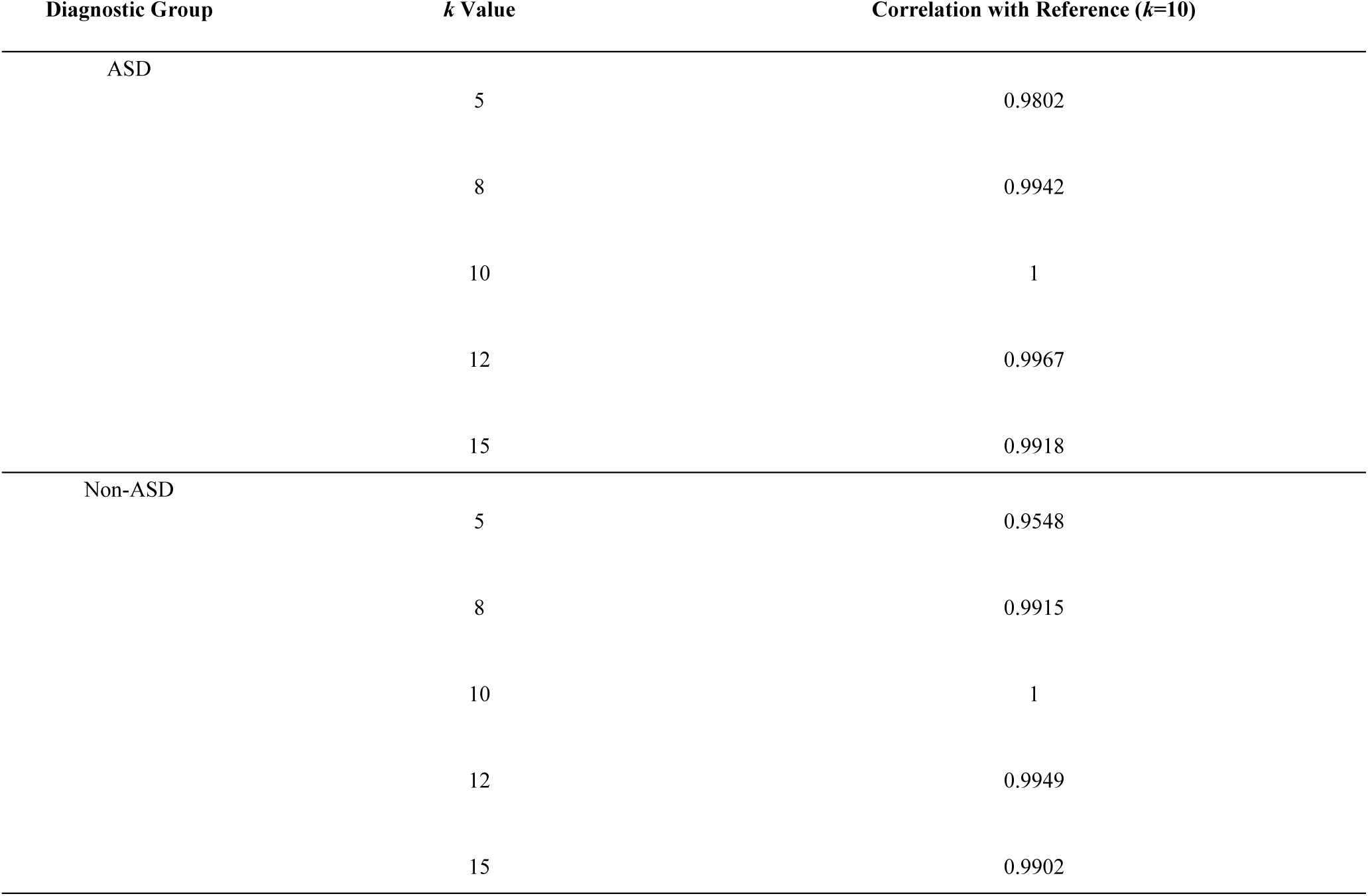
Correlation of the first spectral eigenvector across kNN values. High correlations indicate robustness of the embedding to graph parameters.

| Diagnostic Group | $k$ Value | Correlation with Reference ($k=10$) |
| --- | --- | --- |
| ASD | 5 | 0.9802 |
|  | 8 | 0.9942 |
|  | 10 | 1 |
|  | 12 | 0.9967 |
|  | 15 | 0.9918 |
| Non-ASD | 5 | 0.9548 |
|  | 8 | 0.9915 |
|  | 10 | 1 |
|  | 12 | 0.9949 |
|  | 15 | 0.9902 |

**Table 4:**
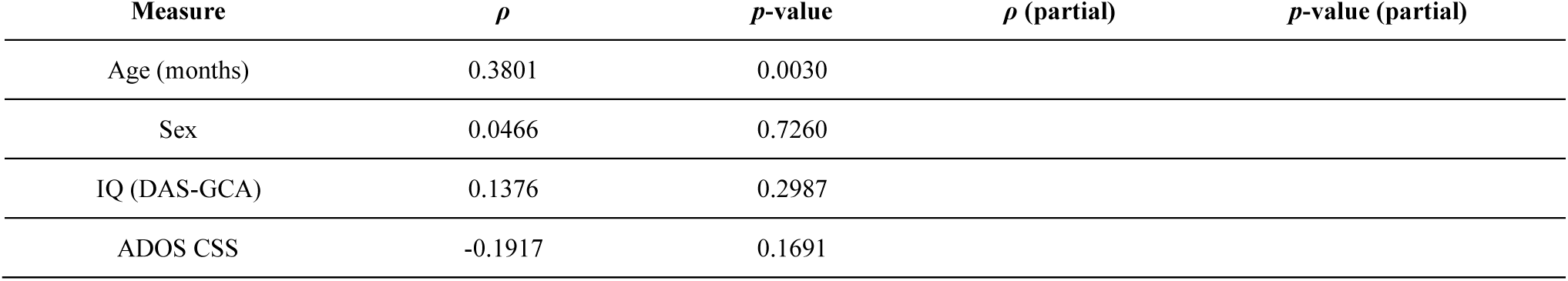
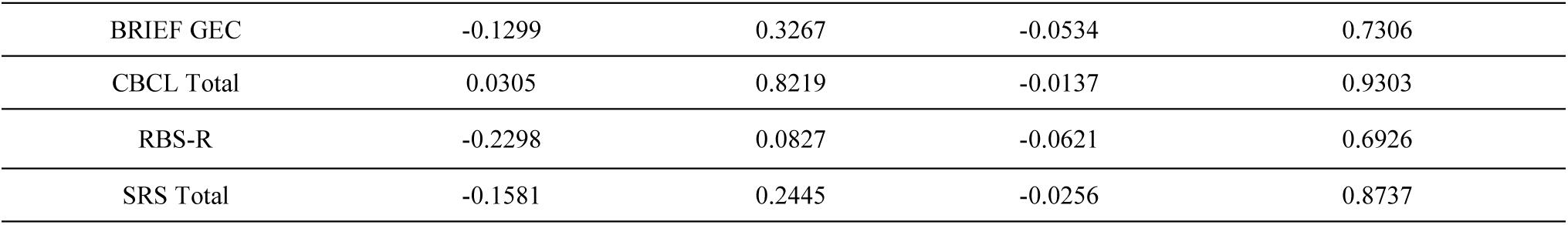
Spearman correlations and partial correlations for the sensory dimension associated with various demographic and behavioral measures. These results reveal no significant association outside of age, likely attributable to modest developmental variation along the sensory axis.

| Measure | $\rho$ | $p$-value | $\rho$ (partial) | $p$-value (partial) |
| --- | --- | --- | --- | --- |
| Age (months) | 0.3801 | 0.0030 |  |  |
| Sex | 0.0466 | 0.7260 |  |  |
| IQ (DAS-GCA) | 0.1376 | 0.2987 |  |  |
| ADOS CSS | -0.1917 | 0.1691 |  |  |
| BRIEF GEC | -0.1299 | 0.3267 | -0.0534 | 0.7306 |
| CBCL Total | 0.0305 | 0.8219 | -0.0137 | 0.9303 |
| RBS-R | -0.2298 | 0.0827 | -0.0621 | 0.6926 |
| SRS Total | -0.1581 | 0.2445 | -0.0256 | 0.8737 |

**Table 5:** Full list of feature importance scores for ASD participant sensory items, ordered by absolute value of Spearman’s *ρ*.

| Rank | Item | Quadrant | Spearman's $\rho$ | Cum. Loading Mass % |
| --- | --- | --- | --- | --- |
| 1 | 25 | Sensory Sensitivity | 0.7220 | 3.5335 |
| 2 | 38 | Sensation Avoiding | 0.6718 | 6.8215 |
| 3 | 22 | Sensory Sensitivity | 0.6457 | 9.9816 |
| 4 | 57 | Sensation Avoiding | 0.6453 | 13.1398 |
| 5 | 51 | Sensory Sensitivity | 0.6408 | 16.2759 |
| 6 | 46 | Sensation Avoiding | 0.6365 | 19.3909 |
| 7 | 54 | Sensory Sensitivity | 0.5937 | 22.2967 |
| 8 | 35 | Sensation Avoiding | 0.5863 | 25.1660 |
| 9 | 56 | Sensation Avoiding | 0.5819 | 28.0140 |
| 10 | 49 | Sensation Avoiding | 0.5634 | 30.7714 |
| 11 | 33 | Sensory Sensitivity | 0.5606 | 33.5149 |
| 12 | 31 | Sensory Sensitivity | 0.5521 | 36.2171 |
| 13 | 24 | Sensation Avoiding | 0.5392 | 38.8559 |
| 14 | 53 | Sensation Avoiding | 0.5176 | 41.3889 |
| 15 | 12 | Low Registration | 0.5138 | 43.9036 |
| 16 | 34 | Sensory Sensitivity | 0.5018 | 46.3596 |
| 17 | 59 | Low Registration | 0.4999 | 48.8063 |
| 18 | 48 | Sensory Sensitivity | 0.4975 | 51.2414 |
| 19 | 37 | Low Registration | 0.4925 | 53.6518 |
| 20 | 45 | Low Registration | 0.4870 | 56.0355 |
| 21 | 16 | Sensory Sensitivity | 0.4820 | 58.3947 |
| 22 | 21 | Low Registration | 0.4666 | 60.6783 |
| 23 | 15 | Low Registration | 0.4589 | 62.9243 |
| 24 | 1 | Sensation Avoiding | 0.4547 | 65.1498 |
| 25 | 52 | Low Registration | 0.4101 | 67.1569 |
| 26 | 13 | Sensory Sensitivity | 0.3938 | 69.0843 |
| 27 | 20 | Sensory Sensitivity | 0.3768 | 70.9286 |
| 28 | 29 | Sensation Avoiding | 0.3691 | 72.7351 |
| 29 | 55 | Low Registration | 0.3460 | 74.4285 |
| 30 | 50 | Sensory Seeking | 0.3003 | 75.8980 |
| 31 | 17 | Sensory Seeking | -0.2999 | 77.3658 |
| 32 | 11 | Sensation Avoiding | 0.2975 | 78.8218 |
| 33 | 60 | Sensory Sensitivity | 0.2923 | 80.2525 |
| 34 | 41 | Low Registration | 0.2885 | 81.6644 |
| 35 | 44 | Low Registration | 0.2816 | 83.0426 |
| 36 | 18 | Sensation Avoiding | 0.2751 | 84.3891 |
| 37 | 27 | Sensory Sensitivity | 0.2680 | 85.7006 |
| 38 | 26 | Sensation Avoiding | 0.2645 | 86.9952 |
| 39 | 8 | Sensory Seeking | 0.2492 | 88.2150 |
| 40 | 5 | Sensation Avoiding | 0.2269 | 89.3258 |
| 41 | 7 | Sensory Sensitivity | 0.2073 | 90.3403 |
| 42 | 4 | Sensory Seeking | -0.1998 | 91.3184 |
| 43 | 6 | Low Registration | 0.1961 | 92.2783 |
| 44 | 23 | Low Registration | 0.1823 | 93.1704 |
| 45 | 58 | Sensory Seeking | -0.1812 | 94.0573 |
| 46 | 2 | Sensory Seeking | 0.1466 | 94.7749 |
| 47 | 9 | Sensory Sensitivity | 0.1442 | 95.4807 |
| 48 | 42 | Sensory Seeking | -0.1279 | 96.1068 |
| 49 | 47 | Sensory Seeking | -0.1214 | 96.7011 |
| 50 | 39 | Low Registration | -0.1091 | 97.2351 |
| 51 | 32 | Sensory Seeking | 0.1006 | 97.7274 |
| 52 | 40 | Sensory Seeking | -0.0980 | 98.2072 |
| 53 | 10 | Sensory Seeking | 0.0913 | 98.6539 |
| 54 | 43 | Sensation Avoiding | 0.0762 | 99.0269 |
| 55 | 3 | Low Registration | 0.0568 | 99.3051 |
| 56 | 14 | Sensory Seeking | -0.0449 | 99.5251 |
| 57 | 30 | Sensory Seeking | -0.0373 | 99.7075 |
| 58 | 36 | Low Registration | -0.0293 | 99.8507 |
| 59 | 28 | Sensory Seeking | 0.0195 | 99.9461 |
| 60 | 19 | Sensory Seeking | -0.0110 | 100.0000 |

**Table 6:** Full list of feature importance scores for non-ASD participant sensory items, ordered by absolute value of Spearman’s *ρ*.

| Rank | Item | Quadrant | Spearman's $\rho$ | Cum. Loading Mass % |
| --- | --- | --- | --- | --- |
| 1 | 52 | Low Registration | 0.5950 | 2.9136 |
| 2 | 59 | Low Registration | 0.5927 | 5.8158 |
| 3 | 51 | Sensory Sensitivity | 0.5778 | 8.6452 |
| 4 | 12 | Low Registration | 0.5728 | 11.4502 |
| 5 | 54 | Sensory Sensitivity | 0.5567 | 14.1760 |
| 6 | 57 | Sensation Avoiding | 0.5544 | 16.8905 |
| 7 | 16 | Sensory Sensitivity | 0.5448 | 19.5580 |
| 8 | 21 | Low Registration | 0.5306 | 22.1563 |
| 9 | 53 | Sensation Avoiding | 0.5293 | 24.7483 |
| 10 | 56 | Sensation Avoiding | 0.5150 | 27.2702 |
| 11 | 48 | Sensory Sensitivity | 0.5053 | 29.7446 |
| 12 | 23 | Low Registration | 0.4921 | 32.1543 |
| 13 | 44 | Low Registration | 0.4825 | 34.5172 |
| 14 | 18 | Sensation Avoiding | 0.4697 | 36.8172 |
| 15 | 15 | Low Registration | 0.4565 | 39.0525 |
| 16 | 25 | Sensory Sensitivity | 0.4512 | 41.2618 |
| 17 | 38 | Sensation Avoiding | 0.4511 | 43.4705 |
| 18 | 45 | Low Registration | 0.4486 | 45.6671 |
| 19 | 22 | Sensory Sensitivity | 0.4418 | 47.8306 |
| 20 | 46 | Sensation Avoiding | 0.4148 | 49.8617 |
| 21 | 55 | Low Registration | 0.4139 | 51.8884 |
| 22 | 39 | Low Registration | 0.4112 | 53.9019 |
| 23 | 20 | Sensory Sensitivity | 0.3740 | 55.7332 |
| 24 | 13 | Sensory Sensitivity | 0.3709 | 57.5493 |
| 25 | 31 | Sensory Sensitivity | 0.3701 | 59.3614 |
| 26 | 37 | Low Registration | 0.3690 | 61.1685 |
| 27 | 4 | Sensory Seeking | 0.3652 | 62.9569 |
| 28 | 24 | Sensation Avoiding | 0.3589 | 64.7144 |
| 29 | 49 | Sensation Avoiding | 0.3494 | 66.4254 |
| 30 | 35 | Sensation Avoiding | 0.3408 | 68.0943 |
| 31 | 1 | Sensation Avoiding | 0.3401 | 69.7597 |
| 32 | 60 | Sensory Sensitivity | 0.3396 | 71.4227 |
| 33 | 29 | Sensation Avoiding | 0.3378 | 73.0769 |
| 34 | 3 | Low Registration | 0.3337 | 74.7111 |
| 35 | 41 | Low Registration | 0.3285 | 76.3194 |
| 36 | 33 | Sensory Sensitivity | 0.3262 | 77.9167 |
| 37 | 36 | Low Registration | 0.3041 | 79.4059 |
| 38 | 43 | Sensation Avoiding | 0.3007 | 80.8785 |
| 39 | 26 | Sensation Avoiding | 0.2980 | 82.3379 |
| 40 | 11 | Sensation Avoiding | 0.2888 | 83.7521 |
| 41 | 50 | Sensory Seeking | 0.2721 | 85.0844 |
| 42 | 6 | Low Registration | 0.2703 | 86.4081 |
| 43 | 2 | Sensory Seeking | 0.2660 | 87.7106 |
| 44 | 34 | Sensory Sensitivity | 0.2644 | 89.0055 |
| 45 | 8 | Sensory Seeking | 0.2208 | 90.0866 |
| 46 | 9 | Sensory Sensitivity | 0.2127 | 91.1279 |
| 47 | 17 | Sensory Seeking | 0.2122 | 92.1671 |
| 48 | 14 | Sensory Seeking | -0.1898 | 93.0965 |
| 49 | 7 | Sensory Sensitivity | 0.1840 | 93.9976 |
| 50 | 5 | Sensation Avoiding | 0.1672 | 94.8164 |
| 51 | 42 | Sensory Seeking | 0.1624 | 95.6114 |
| 52 | 30 | Sensory Seeking | 0.1585 | 96.3876 |
| 53 | 40 | Sensory Seeking | 0.1543 | 97.1433 |
| 54 | 32 | Sensory Seeking | 0.1335 | 97.7972 |
| 55 | 19 | Sensory Seeking | 0.1177 | 98.3737 |
| 56 | 28 | Sensory Seeking | 0.0899 | 98.8141 |
| 57 | 27 | Sensory Sensitivity | 0.0815 | 99.2130 |
| 58 | 58 | Sensory Seeking | 0.0795 | 99.6022 |
| 59 | 47 | Sensory Seeking | -0.0719 | 99.9542 |
| 60 | 10 | Sensory Seeking | 0.0094 | 100.0000 |

**Table 7:**
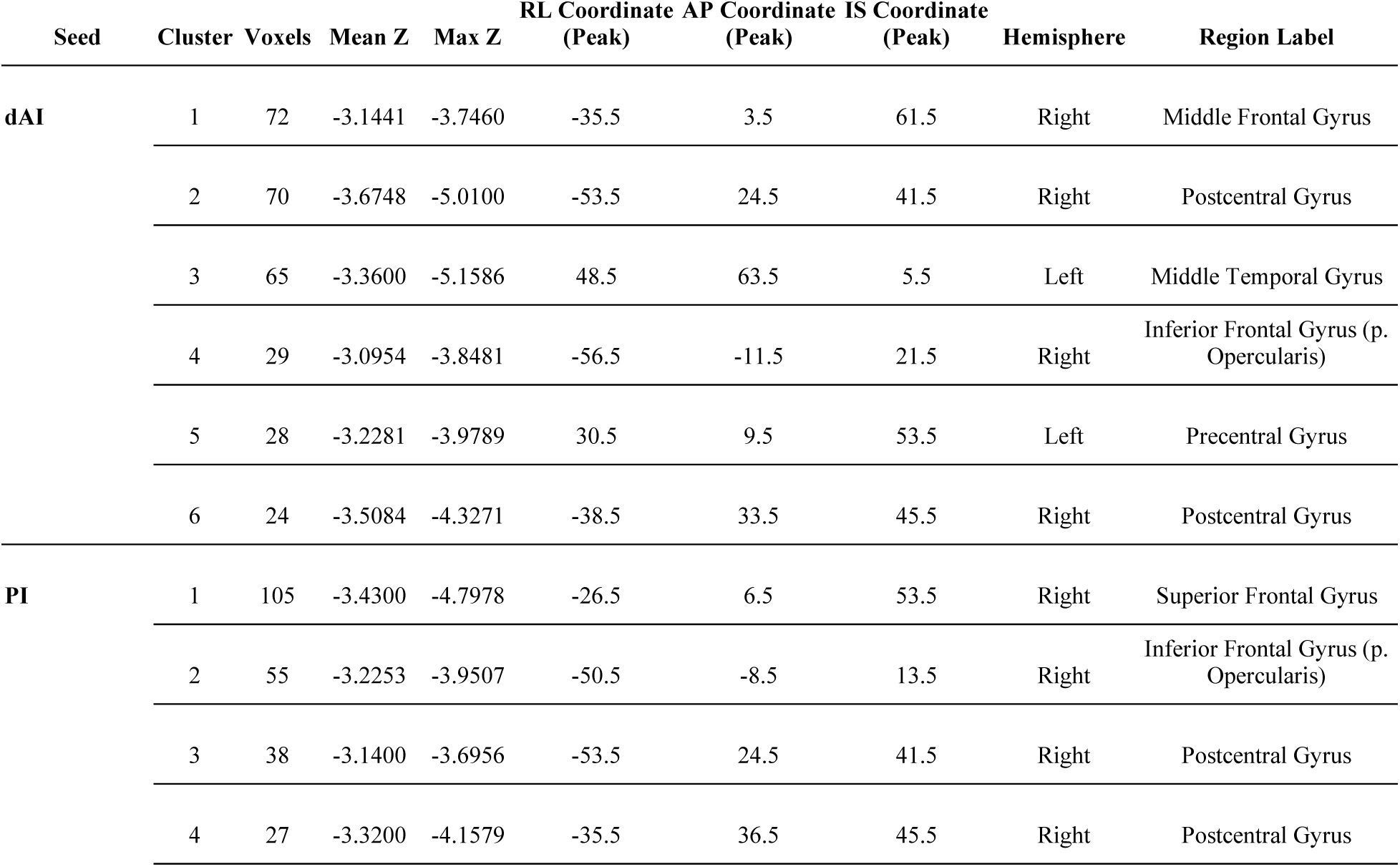
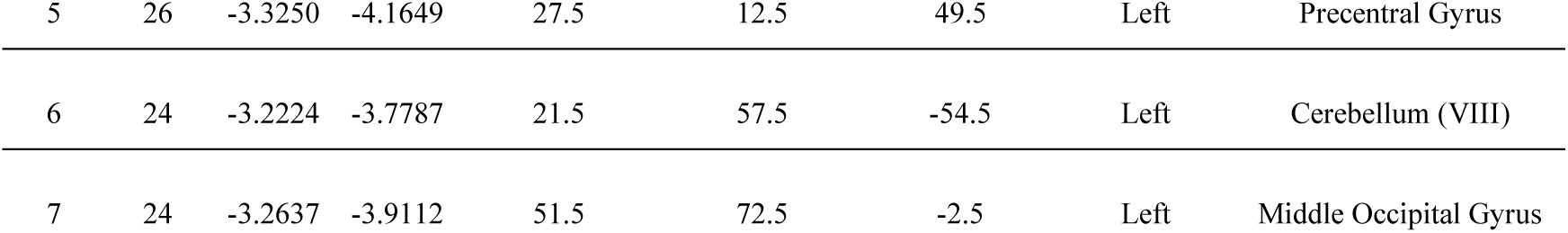
List of significant clusters, mean and maximum z-statistic, peak coordinates, and region labels for both the dorsal anterior insula and posterior insula seeds. RL = right/left, AP = anterior/posterior, IS = inferior/superior.

| Seed | Cluster | Voxels | Mean Z | Max Z | RL Coordinate<br>(Peak) | AP Coordinate<br>(Peak) | IS Coordinate<br>(Peak) | Hemisphere | Region Label |
| --- | --- | --- | --- | --- | --- | --- | --- | --- | --- |
| **dAI** | 1 | 72 | -3.1441 | -3.7460 | -35.5 | 3.5 | 61.5 | Right | Middle Frontal Gyrus |
|  | 2 | 70 | -3.6748 | -5.0100 | -53.5 | 24.5 | 41.5 | Right | Postcentral Gyrus |
|  | 3 | 65 | -3.3600 | -5.1586 | 48.5 | 63.5 | 5.5 | Left | Middle Temporal Gyrus |
|  | 4 | 29 | -3.0954 | -3.8481 | -56.5 | -11.5 | 21.5 | Right | Inferior Frontal Gyrus (p.<br>Opercularis) |
|  | 5 | 28 | -3.2281 | -3.9789 | 30.5 | 9.5 | 53.5 | Left | Precentral Gyrus |
|  | 6 | 24 | -3.5084 | -4.3271 | -38.5 | 33.5 | 45.5 | Right | Postcentral Gyrus |
| **PI** | 1 | 105 | -3.4300 | -4.7978 | -26.5 | 6.5 | 53.5 | Right | Superior Frontal Gyrus |
|  | 2 | 55 | -3.2253 | -3.9507 | -50.5 | -8.5 | 13.5 | Right | Inferior Frontal Gyrus (p.<br>Opercularis) |
|  | 3 | 38 | -3.1400 | -3.6956 | -53.5 | 24.5 | 41.5 | Right | Postcentral Gyrus |
|  | 4 | 27 | -3.3200 | -4.1579 | -35.5 | 36.5 | 45.5 | Right | Postcentral Gyrus |

|  |  |  |  |  |  |  |  |  |
| --- | --- | --- | --- | --- | --- | --- | --- | --- |
| 5 | 26 | -3.3250 | -4.1649 | 27.5 | 12.5 | 49.5 | Left | Precentral Gyrus |
| 6 | 24 | -3.2224 | -3.7787 | 21.5 | 57.5 | -54.5 | Left | Cerebellum (VIII) |
| 7 | 24 | -3.2637 | -3.9112 | 51.5 | 72.5 | -2.5 | Left | Middle Occipital Gyrus |

**Figure 9:**
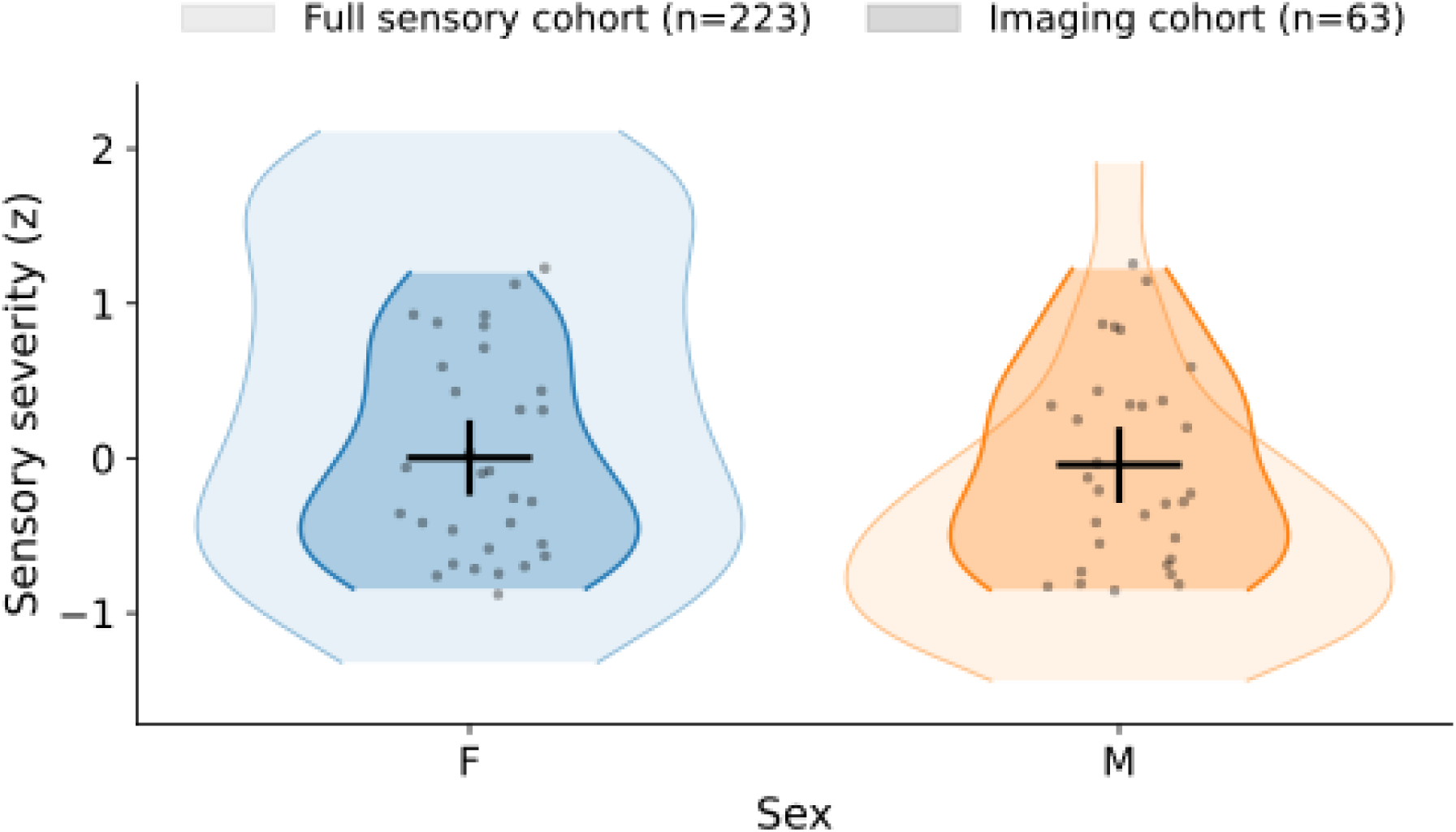
Nested violin plots depict the distribution of the first sensory embedding dimension (standardized to z-scores) separately for females (F) and males (M). The outer, lighter violins represent the full sensory cohort (*n* = 223), while the inner, darker violins with overlaid data points represent the imaging subsample (*n* = 63). Black bars display the mean ±95% confidence interval within the imaging cohort. The imaging subsample spans the central range of the full cohort distribution for both sexes, with no marked sex-based differences in mean sensory severity.

**Table 8:**
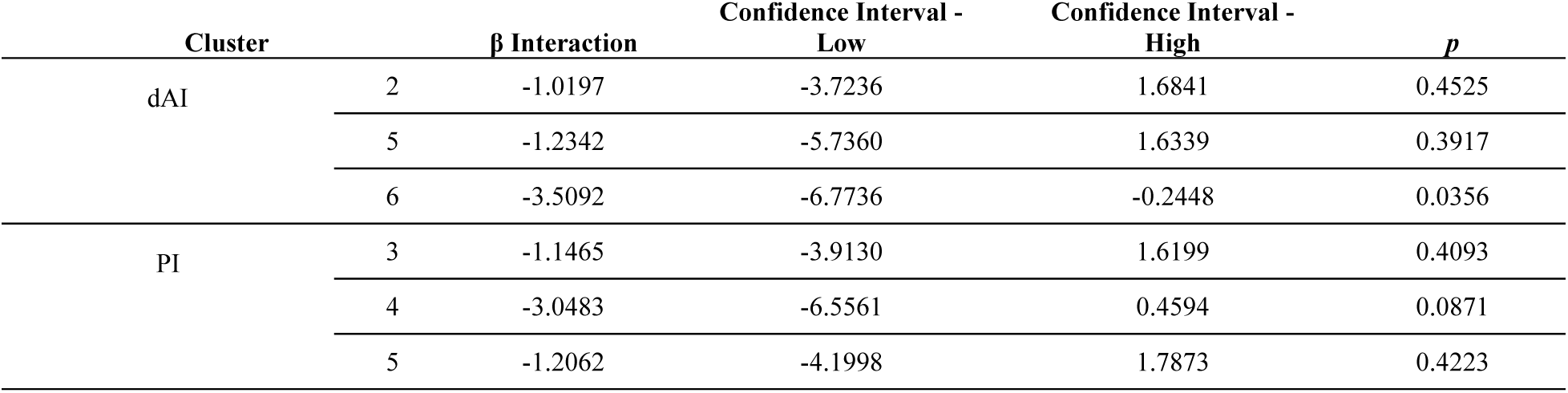
For each cluster, interaction coefficients (β) and 95% confidence intervals are derived from subject-level cluster mean connectivity values using linear models including site, age, IQ, and mean framewise displacement as covariates. Cook’s D values reflect maximum influence statistics across participants (threshold = 4/*n* or 0.0635). The sign flip column indicates whether the interaction coefficient reversed direction under leave-one-out (LOO) resampling. No primary sensorimotor cluster exhibited a sign reversal under LOO validation.

**Table 9:** Interaction coefficients were estimated from subject-level cluster mean connectivity values using linear models including site and relevant covariates. Influence was assessed using Cook’s D (threshold = 4/*n*; 0.0635), and interaction stability was evaluated via leave-one-out (LOO) resampling. The sign flip column indicates whether the interaction coefficient changed direction in any LOO iteration. Clusters emphasized in the main text (sensorimotor regions) exhibited stable interaction direction under LOO validation, whereas peripheral clusters demonstrated greater coefficient variability.

| Cluster | | $\beta$<br>Interaction | Max Cook's<br>D | N above<br>Threshold | LOO Max<br>$\beta$ | LOO Min $\beta$ | LOO Sign<br>Flip | $p$ |
| --- | --- | --- | --- | --- | --- | --- | --- | --- |
| dAI | 1 | -0.5436 | 0.3333 | 6 | 0.1161 | -0.8594 | Yes | 0.6470 |
|  | 2 | -1.0197 | 0.2122 | 5 | -0.0783 | -1.5010 | No | 0.4525 |
|  | 3 | -0.9826 | 0.3000 | 6 | 0.0109 | -1.5954 | Yes | 0.5220 |
|  | 4 | -0.3655 | 0.1309 | 6 | 0.4039 | -0.6589 | Yes | 0.8047 |
|  | 5 | -1.2342 | 0.1327 | 5 | -0.7451 | -1.5919 | No | 0.3917 |
|  | 6 | -3.5092 | 0.4423 | 1 | -1.9449 | -4.2996 | No | 0.0356 |
| PI | 1 | -0.6789 | 0.3082 | 6 | -0.0875 | -0.9517 | No | 0.5590 |
|  | 2 | -0.4737 | 0.1482 | 4 | 0.0827 | -0.7776 | Yes | 0.7188 |
|  | 3 | -1.1465 | 0.2080 | 6 | -0.1930 | -1.6117 | No | 0.4093 |
|  | 4 | -3.0483 | 0.4505 | 1 | -1.3518 | -3.8467 | No | 0.0871 |
|  | 5 | -1.2062 | 0.1742 | 4 | -0.7013 | -1.5379 | No | 0.4223 |
|  | 6 | -0.0981 | 0.1833 | 7 | 0.4975 | -0.6777 | Yes | 0.9304 |
|  | 7 | -1.0765 | 0.2922 | 7 | 0.2585 | -1.6506 | Yes | 0.5636 |

**Table 10:** Comparison between states via paired-samples *t*-test (FD), Wilcoxon signed-rank test (tSNR) and Spearman’s rank correlation between the QC metric and sensory severity. ** *p*<0.05; *** *p*=0.001

| Metric | Biopoint ($M \pm SD$) | Rest ($M \pm SD$) | State Comparison<br>($p$) | Severity $\rho$<br>(Biopoint) | Severity $\rho$ (Rest) |
| --- | --- | --- | --- | --- | --- |
| Mean FD | 0.18±0.17 | 0.30±0.37 | .010** | -0.07 ($p=0.62$) | 0.004 ($p=0.98$) |
| tSNR | 60.6±13.2 | 52.5±16.3 | <0.001*** | 0.01 ($p=0.92$) | -0.03 ($p=0.85$) |

## Acknowledgements

This work was supported by the GENDAAR Consortium (NIH R01 MH100028), and we thank all our colleagues, the participants, and their families for their time and contributions to this research. Members of the GENDAAR Research Consortium who contributed to this work include: James Duncan, PhD; Lawrence Staib, PhD; Lauren Kenworthy, PhD; Raphael Bernier, PhD; Laura Anthony, PhD; Pamela Ventola, PhD; Shefali Jeste, PhD; Michael Crowley, PhD; Charles Nelson, PhD; Daniel Geschwind, PhD; Nicha Dvornek, PhD; Abha Gupta, PhD; Micah Mazurek, PhD; Anna Kresse, MPH; Megha Santhosh, MPA; Emily Neuhaus, PhD; and Jessica Benton, MA.

